# Delays in tuberculosis diagnosis and treatment in India: A patient journey analysis from Mumbai and Patna

**DOI:** 10.1101/2025.10.14.25338008

**Authors:** Mohammad Abdullah Heel Kafi, Poshan Thapa, Madhukar Pai, Charity Oga-Omenka

**Affiliations:** Cardiac Services BC, Provincial Health Service Authority, BC, Canada; Research Institute of the McGill University Health Centre, Montréal, Quebec, Canada; McGill International TB Centre, Montreal, Canada; Department of Epidemiology, Biostatistics and Occupational Health, McGill University, Montreal, Canada; School of Public Health Sciences, University of Waterloo, Waterloo, Canada

## Abstract

**Background:** Tuberculosis (TB) patient services in India are often fragmented, undermining timely access to timely diagnosis and treatment. Understanding patient journeys is critical to strengthening TB care delivery and achieving elimination goals.

**Methods:** We conducted a cross-sectional study of 400 TB patients diagnosed between 2020– 2022 in two major Indian cities: Mumbai (n=200) and Patna (n=200). Using structured interviews, we examined health-seeking behavior, delays to diagnosis and treatment, the number and type of healthcare encounters, and out-of-pocket costs.

**Results:** Patients predominantly initiated care in the private sector (91% in Mumbai; 85% in Patna), often with pharmacies or private clinics. Care pathways were fragmented, requiring multiple provider visits before diagnosis. The median total delay from symptom onset to treatment initiation was 35 days (IQR: 13–81) in Patna and 26 days (IQR: 12–59) in Mumbai. Provider delays accounted for nearly 19 days in both settings. Patients made a median of 3 healthcare visits pre-diagnosis, with 23% experiencing ≥6 encounters. The financial burden of TB care was substantial, particularly in Mumbai, where consultation and diagnostic costs were markedly higher than in Patna. Longer delays and higher numbers of encounters were associated with being male, unemployed, having larger household size, and hesitation to seek care during the study period.

**Conclusion:** TB patient pathways in urban India pandemic were prolonged, costly, and fragmented — especially within the private sector during the COVID-19. Strengthening public-private integration, improving early diagnosis strategies, and protecting patients from financial hardship are essential priorities to accelerate TB elimination and strengthen health system resilience against future disruptions.

## INTRODUCTION

Tuberculosis (TB), an infectious disease caused by Mycobacterium tuberculosis, remains a significant public health issue, with India accounting for 26% of the global TB burden[1]. In 2022, approximately 2.77 million TB cases were reported in the country [1,2]. However, undiagnosed or “missing” TB cases remain a significant challenge, with up to 25% of cases unreported, further adding to transmission and disease burden [3]. The slow decline in TB incidence rates is primarily attributed to persistent socio-economic factors such as poverty, malnutrition, overcrowded living conditions, and limited access to healthcare services.

Compounding these challenges is a large, underregulated, and diverse private sector, which ranges from small clinics to multi-specialty hospitals and includes informal healthcare providers and highly qualified specialists, catering to up to 50% of TB patients in India [4–7]. Furthermore, health system disruptions can reveal critical vulnerabilities in TB care delivery, particularly in densely populated countries like India.

This study examines the COVID-19 pandemic serves as a case study of health system disruptions, with lessons that remain relevant for strengthening TB services against future crises. The pandemic exacerbated pre-existing challenges, severely disrupting TB treatment services, case detection, and reporting processes [8,9]. National lockdowns, social distancing measures, and the diversion of healthcare resources towards COVID-19 management significantly reduced the availability of diagnostic and treatment services for TB [10–12]. For instance, TB case notifications in India dropped by 25% in 2020 compared to the previous year, with the most substantial decline observed during the nationwide lockdown from March to April [13]. This decline in case detection corresponded with an 11% increase in TB-related mortality in 2020 compared to 2019 [14]. These disruptions were even more pronounced in India’s private healthcare sector, which accounts for an estimated 60-85% of initial TB care interactions occur, resulting in a 44% decline in case notifications during the lockdown period [11].

Understanding the systemic weaknesses in TB care revealed during the pandemic requires a comprehensive examination of patients’ healthcare-seeking behaviors and their pathways to diagnosis and treatment. In this context, Patient Journey Analyses (PJAs) offer a valuable approach for identifying delays in diagnosis, treatment, and healthcare visits, as well as exploring systemic barriers that hinder access to timely care [15–18]. By mapping patients’ healthcare journeys, PJAs help uncover missed opportunities for early diagnosis and timely treatment initiation and highlight gaps across public and private healthcare sectors. This analytical approach is particularly valuable for identifying persistent vulnerabilities even as health systems recover from major disruptions. However, conventional PPAs have notable limitations, including their inability to measure the duration of delays, their tendency to simplify complex patient trajectories, and their limited capacity to capture individual-level factors associated with prolonged pathways. These challenges have been further compounded by the COVID-19 pandemic, which disrupted healthcare systems and added new layers of complexity to patients’ access to TB services, effects that remain insufficiently studied.

Despite the well-documented impact of COVID-19 on TB case detection, there remains a paucity of research exploring the specific delays and barriers encountered by TB patients during the pandemic, particularly in densely populated urban centers in India. While previous studies have primarily focused on aggregate changes in TB notification rates [19,20]. They lack a granular analysis of patient experiences or healthcare interactions, contributing to delays. By focusing on the patient pathway, this study addresses a critical gap in the literature by exploring how system-level shocks affect individual patient journeys, with implications for ongoing TB control efforts and future emergency preparedness.

Mumbai and Patna were purposefully selected for this study to capture contrasting healthcare situations in India during the COVID-19 pandemic. Mumbai, one of the most densely populated cities globally, reported over 85,000 COVID-19 cases by mid-2020 and became an early epicentre of the pandemic in India, resulting in significant strain on its healthcare infrastructure [21]. Mumbai also has a high burden of TB, including drug-resistant TB, particularly in overcrowded settlements such as Dharavi [22]. The pandemic compounded these challenges, with studies showing a 28% decline in TB case notifications between March 2020 and March 2021 and a 69% decline during the strict lockdown period alone [23] . In contrast, Patna, a major city in Bihar, represents a setting with weaker baseline healthcare infrastructure and longstanding barriers to healthcare access, which were further exacerbated during COVID-19 by widespread disruptions and reverse migration from urban to rural areas [24]. Bihar’s TB services were notably affected, reflecting broader national trends of an 80% reduction in TB notifications during lockdown phases [25].

By examining patient pathways in these two settings, this study aims to uncover delays in diagnosis and treatment, the frequency of healthcare interactions required for diagnosis, and the factors associated with delays in care during the COVID-19 pandemic. These findings provide evidence to inform approaches to building more resilient health system, highlighting where targeted interventions can strengthen TB care against existing challenges and future disruptions.

## METHODS

### Study Design and Setting

This cross-sectional study was conducted in Mumbai and Patna, India, between February and March 2022. The study was part of a larger mixed-methods study aimed at identifying disruptions to TB care during the COVID-19 pandemic and identifying health system adaptations that could strengthen health system resilience. Findings from this broader study have already been reported in several publications [26–29]. This manuscript focuses on the quantitative aspects of a broader study, specifically examining tuberculosis (TB) diagnosis and treatment delays, as well as the factors contributing to these delays during the COVID-19 pandemic. A companion qualitative analysis providing in-depth patient narratives (n=39) from the same study sites is reported separately [30]. The study in India was facilitated by the Institute for Social and Economic Research on Development and Democracy (ISERDD), a research organization with an established presence in urban health across multiple regions of India, including Mumbai and Patna.

### Study Participants and Sampling

This study included 400 participants, 200 each recruited from each city (Mumbai and Patna), who were purposively selected to reflect a range of care-seeking experiences during the COVID-19 pandemic. Participants were eligible if they were 18 years old or older and had sought healthcare services for symptoms suggestive of TB or had been diagnosed with TB during the COVID-19 pandemic period. Study recruitment focused on patients diagnosed with TB during the COVID-19 pandemic (2020-2022). A purposive sampling strategy was employed to ensure participants represented diverse healthcare interactions, including those who accessed care from both the public and private sectors. Recruitment was conducted by trained field officers affiliated with ISERDD, who also had prior experience with community-based TB research.

### Outcomes of Interest and Definitions

The primary outcomes were: 1) delays in TB diagnosis and treatment initiation, and 2) the type and number of healthcare encounters involved in the patient’s care-seeking pathway. Fig 1 outlines the different types of delays: (i) Health-seeking delay is defined as the number of days from the onset of symptoms to the first healthcare consultation; (ii) Provider delay is defined as the number of days from the first consultation to TB diagnosis, and (iii) Treatment delay is defined as the number of days from diagnosis to treatment initiation [31,32]. An encounter was defined as any interaction with a healthcare provider, including visits to pharmacists, clinics, hospitals, informal providers, and specialists. The study also assessed the frequency and patterns of healthcare visits as well as the types of healthcare facilities assessed, including private clinics, public hospitals, and pharmacies.

**Fig 1.**
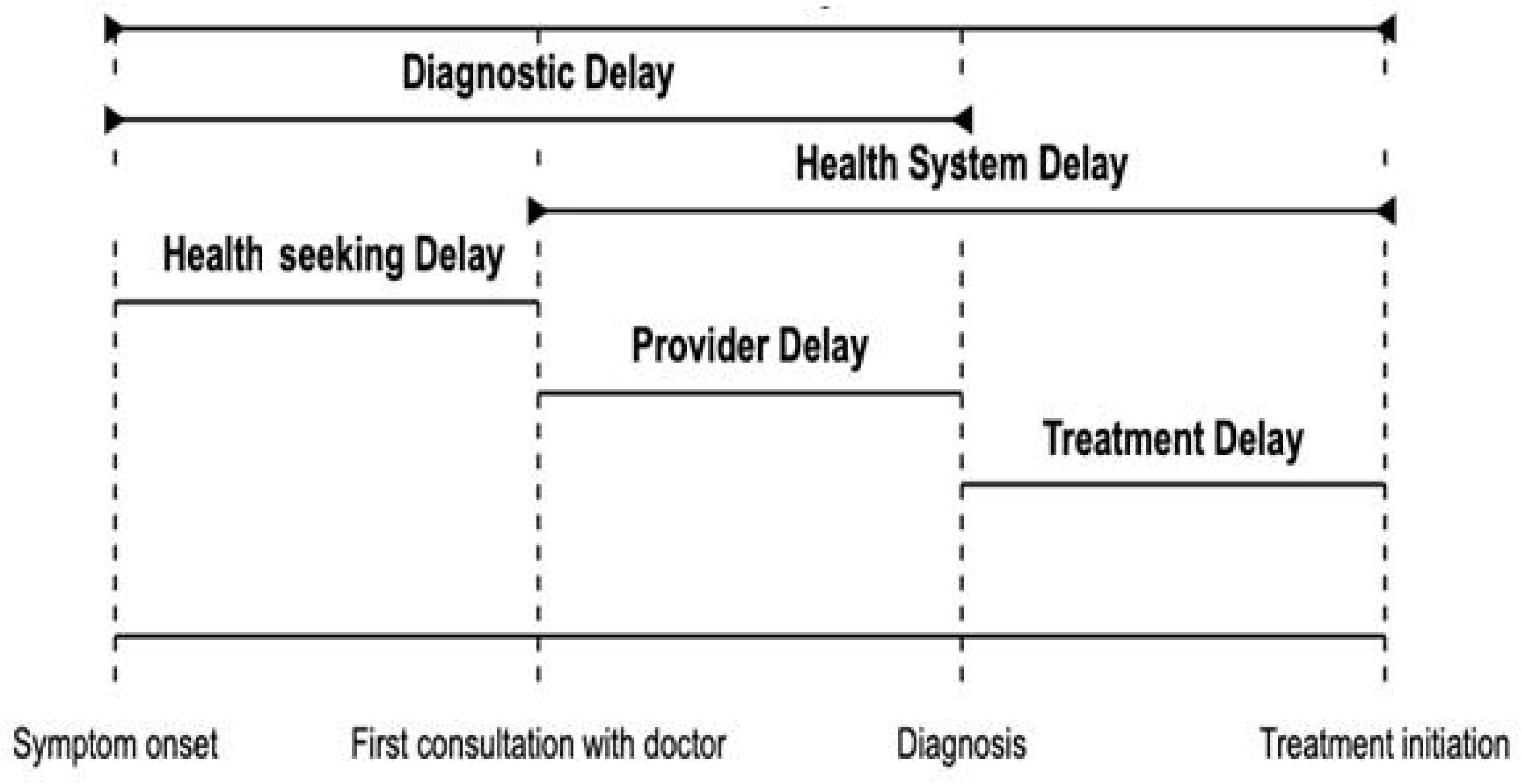
Definition of delays in healthcare-seeking, diagnosis, and treatment, figure recreated for clarity, adapted from the World Health Organisation.

This study collected all delay measurements through structured patient interviews using a standardized questionnaire administered by trained field interviewers. Patients were asked to recall the dates when their symptoms began, their first healthcare contact, and when TB diagnosis and treatment were initiated. Interviewers used calendar prompts to improve recall accuracy for patients who were unable to recall exact dates. For encounter information from the patient, all healthcare visits between symptom onset and treatment initiation were documented, including the type of provider, diagnostic tests performed, and recommendations received.

A cut-off of 30 days was used to define a health-seeking delay, and a cut-off of six encounters was used to indicate a high number of healthcare interactions, based on the same set of predictors as described above. The rationale for these cut-offs was derived from established clinical guidelines and prior studies on TB care pathways [33]. These thresholds were used in subsequent analyses of factors associated with delays in the TB care pathway.

### Study tool and data collection

The data collection tool was developed based on a review of previous studies on the TB patient journey and was adapted to include questions specific to the COVID-19 context and disruptions. The structured questionnaire contained sections on socio-demographic characteristics, symptom history, healthcare-seeking behavior, diagnostic experiences, treatment initiation, and COVID-19-related barriers and disruptions. The tool was translated into the local language (Hindi) and then back translated to ensure linguistic accuracy. It was pre-tested with a few TB patients not included in the final sample, which led to refinements in question phrasing and response categories. The final version of the questionnaire is provided in the supporting information (Table A in S1 Text).

A team of trained research assistants with public health backgrounds and prior TB research experience conducted data collection. All data collectors received training on study procedures, questionnaire administration, and research ethics. Interviews were conducted in-person at TB treatment centers using tablet-based electronic forms. The study coordinator performed weekly quality checks to identify and address data inconsistencies or missing information. To minimize recall bias, interviewers used calendar tools and referred to significant personal and national events to help patients accurately recall the dates and details of their healthcare interactions.

### Statistical Analysis

All data cleaning, visualization, and statistical analysis were performed using statistical software R version 4.4.1 [34]. Numerical variables were described by median and interquartile range (IQR), and categorical variables were described using proportion (percentage). Due to the skewed distribution of continuous outcomes and the presence of outliers, quantile regression (tau = 0.5) was employed to investigate the relationship between the outcome variables (various delays) and potential predictors. Linear regression was used to analyze the number of healthcare encounters.

Univariate regression was initially used to explore factors associated with delays and the number of encounters. Multivariate models were then fitted to control for confounders and to examine the impact of COVID-19 on the outcomes of interest. Variables considered in the multivariate analysis were selected based on previous literature [33] and systematic reviews summarising risk factors for delays in TB care [35–38], which included age, gender, marital status, household size, education level, employment status, and financial standing, as well as clinical and exposure-related factors such as contact history with active TB cases, having friends or family with COVID-19, and whether COVID-19 affected the willingness to seek healthcare. Additional factors, such as distance to the nearest public and private healthcare facilities to explore geographic accessibility in delays [35–38], as well as internet usage history, which may reflect differential access to healthcare information during the pandemic, were included in the outcome related to the number of encounters. Sensitivity analyses were conducted using logistic regression models to validate findings from the quantile regression and assess the robustness of associations. A cut-off of 30 days was used to define health-seeking delay, and a cut-off of six encounters was used to indicate a high number of healthcare interactions, with all the same predictors as mentioned above. The rationale for these cut-offs was derived from established clinical guidelines and prior studies on TB care pathways [33] These thresholds were used for subsequent analyses of factors associated with delays in the TB care pathway.

### Ethical consideration

This study was funded by the Bill and Melinda Gates Foundation (INV022420). Ethical approval was obtained from the McGill University Research Ethics Board (Covid BMGF / 2021-7197), the Georgetown-Medstar IRB (STUDY00003422), and the Institute for Social and Economic Research on Development and Democracy (ISERDD). The funders had no role in the study design, data collection and analysis, the decision to publish, or the preparation of the manuscript. The recruitment process ensured that participants provided informed consent before data collection.

## RESULTS

### Participants Baseline Characteristics

The baseline characteristics of the study participants from Patna and Mumbai, presented in Table 1, showed distinct demographic and healthcare access patterns in both cities. In Patna, the mean age of participants was 39.1 years, with 60% being male; in Mumbai, the mean age was 31.6 years, with 63% being female. Marital status also varied, with a higher proportion of single individuals in Mumbai (43.5%) compared to Patna (30%). Participants in Patna reported slightly larger household sizes (mean = 4.68, SD = 2.44) than those in Mumbai (mean = 4.09, SD = 2.31). Educational attainment was notably higher among Mumbai participants, with 23% having completed a bachelor’s degree, compared to 14.5% in Patna. Employment status was similar across both cities, with more than half of participants being unemployed or underemployed (52% in Patna and 54.5% in Mumbai). Financial adequacy was reported by approximately half of the participants in both Patna (46.5%) and Mumbai (53.5%).

**Table 1.** Baseline characteristics of the study sample (N = 400).

| **Characteristics** | **Patna (N =200)** | **Mumbai (N =200)** |
| --- | --- | --- |
| **Age** |  |  |
| Mean (SD) | 39.1(15.9) | 31.6 (11.0) |
| Missing |  | 6 (3.0%) |
| **Gender** |  |  |
| Male | 120 (60%) | 70 (35.0%) |
| Female | 80 (40%) | 126 (63.0%) |
| Missing |  | 4 (2.0%) |
| **Marital status** |  |  |
| Single | 60 (30%) | 87 (43.5%) |
| Married | 118 (59%) | 104 (52.0%) |
| Widowed | 22 (11%) | 0 (0%) |
| Missing |  | 4 (2.0%) |
| **Individuals in house** |  |  |
| Mean (SD) | 4.68 (2.44) | 4.09 (2.31) |
| Missing |  | 4 (2.0%) |
| **Highest education level** |  |  |
| Primary 6 or lower | 60 (30%) | 31 (15.5%) |
| Junior secondary school | 49 (24.5%) | 50 (25.0%) |
| Senior secondary school | 18 (9%) | 51 (25.5%) |
| Bachelor's degree | 29 (14.5%) | 46 (23.0%) |
| Post-graduate certificate | 9 (4.5%) | 10 (5.0%) |
| Other | 33 (16.5%) | 7 (3.5%) |
| Missing |  | 4 (2.0%) |
| **Employment** |  |  |
| Working | 60 (30%) | 80 (40.0%) |
| Self-employed | 30 (15%) | 6 (3.0%) |
| Unemployed/underemployed | 104 (52%) | 109 (54.5%) |
| Missing | 6 (3%) | 4 (2.0%) |
| **Financial standing** |  |  |
| Adequate | 93 (46.5%) | 107 (53.5%) |
| Barely adequate | 72 (36%) | 73 (36.5%) |
| Inadequate | 35 (17.5%) | 15 (7.5%) |
| Missing |  | 5 (2.5%) |

### Healthcare Access and TB Status

Most participants in both cities had good access to healthcare facilities, with 63.5% of participants in Patna and 79.5% in Mumbai living within 2 km of a public healthcare center. Similarly, most participants in both cities reported proximity to private health centers, with 96.5% of participants in Patna and 94.5% in Mumbai residing within 2 km of a private facility (Table 2).

**Table 2.** Distribution of study participants based on TB-related characteristics and healthcare Access.

| **Characteristics** | **Patna (N =200)** | **Mumbai (N =200)** |
| --- | --- | --- |
| **Nearest public health center** |  |  |
| Less than 2 km away | 127 (63.5%) | 159 (79.5%) |
| Between 2-10 km away | 71 (35.5%) | 32 (16.0%) |
| Over 10km | 2 (1%) | 2 (1.0%) |
| Don't know/not sure |  | 2 (1.0%) |
| Missing |  | 5 (2.5%) |
| **Nearest private health center** |  |  |
| Less than 2 km away | 193 (96.5%) | 189 (94.5%) |
| Between 2-10 km away | 7 (3.5%) | 5 (2.5%) |
| Over 10km |  | 0 (0%) |
| Missing |  | 6 (3.0%) |
| **Symptoms that prompted a visit to a healthcare provider<sup>a</sup>** |  |  |
| Cough | 175 (87.5%) | 121 (60.5%) |
| Chest congestion | 93 (46.5%) | 39 (19.5%) |
| Head congestion | 50 (25.0%) | 20 (10.0%) |
| Sore throat | 72 (36.0%) | 18 (9.0%) |
| Rashes or allergies | 14 (7.0%) | 3 (1.5%) |
| Trouble breathing/wheezing | 90 (45.0%) | 44 (22.0%) |
| Weight loss | 154 (77.0%) | 92 (46.0%) |
| Night sweats | 59 (29.5%) | 15 (7.5%) |
| Fever | 155 (77.5%) | 124 (62.0%) |
| Lack of appetite | 143 (71.5%) | 85 (42.5%) |
| **TB care stage now** |  |  |
| Currently taking TB medication | 76 (38.0%) | 110 (55.0%) |
| Completed all TB medication | 108 (54.0%) | 85 (42.5%) |
| **Location of TB diagnosis** |  |  |
| Public Hospital | 22 (11.0%) | 2 (1.0%) |
| Private outpatient clinic, health center, or dispensary | 172 (86.0%) | 86 (43.0%) |
| Private hospital or nursing home | 5 (2.5%) | 107 (53.5%) |
| **Diagnosis is given in the same location as the first encounter** |  |  |
| No | 105 (52.5%) | 144 (72.0%) |
| Yes | 95 (47.5%) | 52 (26.0%) |
| Missing |  | 4 (2.0%) |
| **Location of TB Treatment** |  |  |
| Community health worker/CHEW | 1 (0.5%) | 2 (1.0%) |
| Public Hospital | 24 (12.0%) | 5 (2.5%) |
| Private outpatient clinic, health center, or dispensary | 170 (85.0%) | 60 (30.0%) |
| Private hospital or nursing home | 5 (2.5%) | 129 (64.5%) |
| **Treatment is given in the same location as the site of diagnosis** |  |  |
| No | 53 (26.5%) | 76 (38.0%) |
| Yes | 147 (73.5%) | 120 (60.0%) |
| Missing |  | 4 (2.0%) |

<sup>a</sup> Patients could select multiple symptoms; percentages do not sum to 100%.

Symptoms prompting healthcare visits differed between the two cities, with cough, weight loss, and fever being more frequently reported in Patna (87.5%, 77%, and 77.5%, respectively) compared to Mumbai (60.5%, 46%, and 62%). A higher proportion of participants in Mumbai were still taking TB medication at the time of the survey (55%) compared to Patna (38%). In Patna, 86% of participants were diagnosed at private outpatient clinics, while 53.5% of participants in Mumbai were diagnosed at private hospitals or nursing homes. Similarly, treatment was predominantly administered at private outpatient clinics in Patna (85%), whereas 64.5% of participants in Mumbai received treatment at private hospitals or nursing homes. More details of the symptoms that led patients to seek care from various providers are presented in the supporting information (Table B in S1 Text).

### Distribution of Delays During COVID-19

The distribution of delays experienced by TB patients during the COVID-19 pandemic in Patna and Mumbai is detailed in Fig 2. The median health-seeking delay was 0 days in Patna (IQR 0 to 5 days) compared to 3 days in Mumbai (IQR 2 to 6 days). Provider delays showed similarity in both cities, with 19.5 days in Patna (IQR 4 to 57 days) and 18.5 days in Mumbai (IQR 7 to 43 days). However, diagnostic delays were notably longer in Patna, with a median of 31 days (IQR, 9-68), compared to 23 days in Mumbai (IQR, 10 to 54 days). Treatment delays were minimal in both cities, with a median of 1 day suggesting that treatment initiation was relatively prompt following a diagnosis. Health system delays, which encompass cumulative delays within the healthcare system, had a median of 27 days in Patna (IQR, 7 to 74 days) and 22 days in Mumbai (IQR, 9 to 51 days). The total delay, from initial health-seeking to treatment initiation, was longer in Patna, with a median of 35 days (IQR, 13 to 81 days), compared to 26 days in Mumbai (IQR, 12 to 59 days).

**Fig 2.**
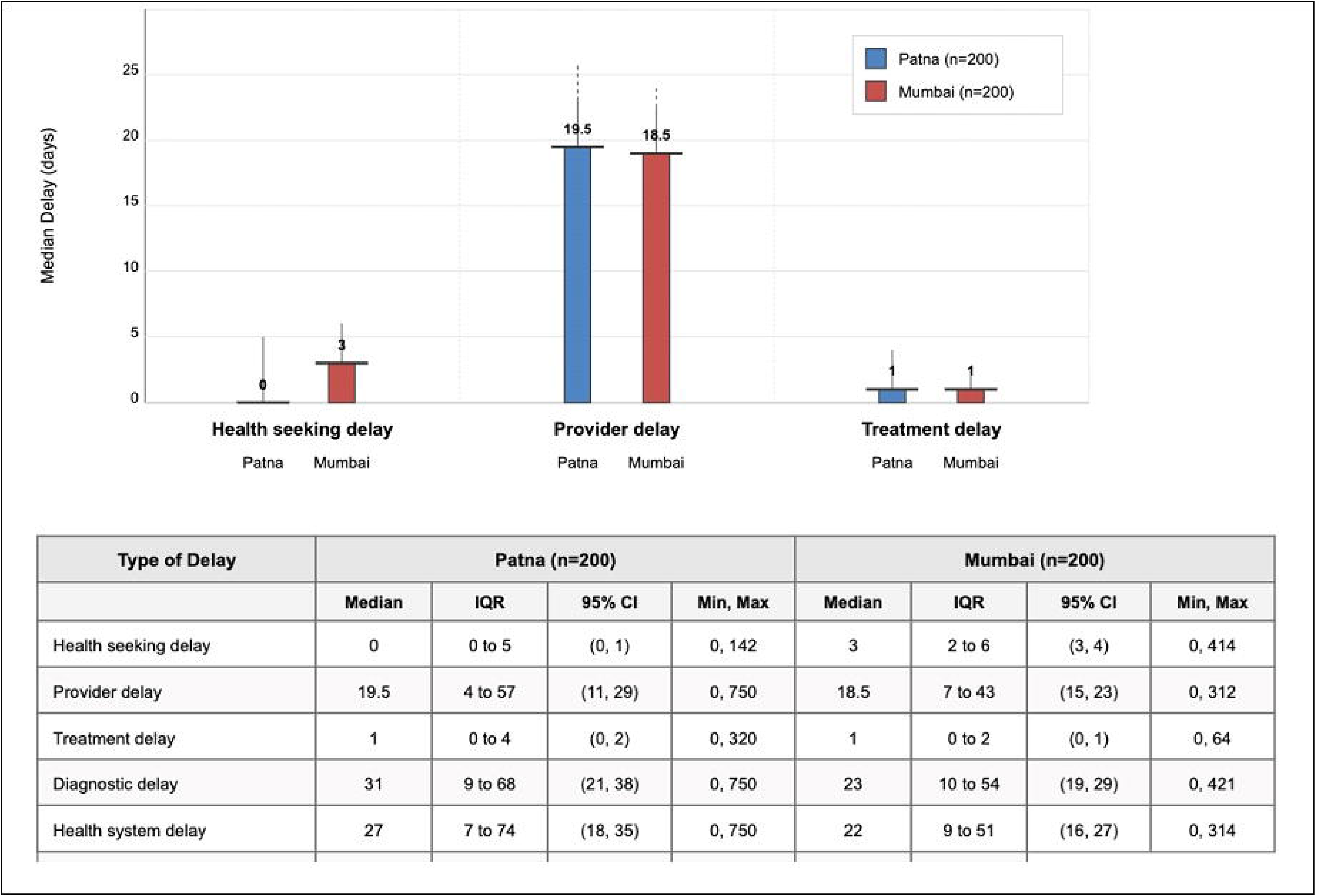
TB Diagnostic and Health System Delays During COVID-19 in Patna and Mumbai.

### Impact of COVID-19 on TB Care Utilization and Associated Cost

The impact of the COVID-19 on TB healthcare and utilization exposed distinct challenges in Patna and Mumbai, as presented in Table 3. Healthcare access disparities were particularly pronounced, with 14.5% of participants in Patna unable to reach a doctor due to facility closures, compared to just 4% in Mumbai. Similarly, medication access was disrupted for 4% of participants in Patna and only 0.5% in Mumbai. Movement restrictions further hindered access, preventing 12% of participants in Patna and 8% of participants in Mumbai from seeking care. However, prolonged waiting times were more commonly reported in Mumbai (21.5%) than in Patna (9.5%).

Behavioral changes related to healthcare-seeking were more pronounced in Mumbai, where 50.5% of participants reported an increased willingness to seek healthcare, largely due to early recognition of symptoms (50%). In Patna, 31.5% of participants reported an increased willingness to seek care.

The adaptability of the healthcare system was evident, with private outpatient clinics serving as the primary source of TB care in both cities (90.5% in Patna and 91% in Mumbai). However, there was a stark contrast in the use of private hospitals, with 60% of participants in Mumbai utilizing private hospitals or nursing homes, compared to only 3% in Patna. Unmet healthcare needs were reported by 9.5% of participants in Patna, compared to 2.5% in Mumbai, with facility closure being the primary reason for unmet needs. Despite the increased need for alternative healthcare service options during the pandemic, telemedicine use remained minimal, with only slight increases observed in both cities (5.5% in Patna and 2.5% in Mumbai).

**Table 3.** The Impact of the COVID-19 Pandemic on TB Healthcare Utilization and Cost.

| Characteristic | Patna<br>(N=200) | Mumbai<br>(N=200) |
| --- | --- | --- |
| **Perspectives on COVID-19 and Healthcare Access** |  |  |
| **Healthcare access impacts** |  |  |
| Unable to reach a doctor because facilities were closed | 29 (14.5%) | 8 (4.0%) |
| Unable to receive medication because drug stores were closed | 8 (4.0%) | 1 (0.5%) |
| Obligated to seek care from a different facility than preferred | 7 (3.5%) | 3 (1.5%) |
| Waiting time became longer than normal/expected | 19 (9.5%) | 43 (21.5%) |
| Unable to leave the house to seek care due to movement restrictions | 24 (12.0%) | 16 (8.0%) |
| Other effects on healthcare | 4 (2.0%) | 132 (66.0%) |
| **Friends/family members diagnosed with COVID** |  |  |
| Yes | 25 (12.5%) | 33 (16.5%) |
| **Willingness to seek healthcare** |  |  |
| More willing | 63 (31.5%) | 101 (50.5%) |
| Less willing | 19 (9.5%) | 10 (5.0%) |
| No effect | 118 (59.0%) | 83 (41.5%) |
| **Reasons for increased willingness** |  |  |
| Early healthcare seeking | 58 (29.0%) | 100 (50.0%) |
| Awareness of family member's infection | 26 (13.0%) | 6 (3.0%) |
| Awareness of getting infected by others | 17 (8.5%) | 6 (3.0%) |
| Understanding of underlying/chronic condition | 17 (8.5%) | - |
| **Healthcare Utilization Patterns** |  |  |
| **Types of contacted health providers <sup>a</sup>** |  |  |
| Community pharmacy, chemist, drug shop, patent medicine vendor | 105 (52.5%) | 3 (1.5%) |
| Private laboratory | 34 (17.0%) | 1 (0.5%) |
| Public outpatient clinic (e.g., PHC/Health Post/Dispensary) | 5 (2.5%) | 5 (2.5%) |
| Public Hospital | 59 (29.5%) | 44 (22.0%) |
| Private outpatient clinic, health center, or dispensary | 181 (90.5%) | 182 (91.0%) |
| Private hospital or nursing home | 6 (3.0%) | 120 (60.0%) |
| **First source for treatment seeking** |  |  |
| Community pharmacy, chemist, drug shop, patent medicine vendor | 82 (41.0%) | - |
| Public Hospital | 12 (6.0%) | 2 (1.0%) |
| Private outpatient clinic, health center, or dispensary | 103 (51.5%) | 184 (92.0%) |
| Private hospital or nursing home | 2 (1.0%) | 5 (2.5%) |
| **Unmet healthcare needs** |  |  |
| No | 181 (90.5%) | 191 (95.5%) |
| Yes | 19 (9.5%) | 5 (2.5%) |
| **Reason for unmet healthcare needs** |  |  |
| Healthcare was not available because the practice was closed due to COVID-19 | 13 (6.5%) | 3 (1.5%) |
| **Telemedicine use before the pandemic** |  |  |
| Did not use telemedicine | 191 (95.5%) | 194 (97.0%) |
| Telemedicine use since pandemic start |  |  |
| Did not use telemedicine | 181 (90.5%) | 188 (94.0%) |
| Once a month or less | 11 (5.5%) | 5 (2.5%) |
| **Healthcare Costs** |  |  |
| Current healthcare costs compared to before COVID-19 |  |  |
| Costs stayed the same | 50 (25.0%) | 19 (9.5%) |
| Costs increased | 127 (63.5%) | 171 (85.5%) |
| **Consultation fee at this facility (Indian rupees)** |  |  |
| Average fee (SD) | 326 (237) | 564 (461) |
| **Lab test price at this facility (Indian rupees)** |  |  |
| Average price (SD) | 2550 (3110) | 9970 (12600) |
| **Monthly TB medication price (Indian rupees)** |  |  |
| Average price (SD) | 188 (660) | 114 (372) |

<sup>a</sup> Patients could go multiple providers; percentages do not sum to 100%.

A significant proportion of patients in both cities experienced increased healthcare costs, with 63.5% of participants in Patna and 85.5% in Mumbai reported higher expenses compared to pre-pandemic levels. Healthcare expenses in Mumbai were notably higher across various categories than in Patna. The average consultation fee in Mumbai was 564 Indian Rupees (SD = 461), nearly double that of Patna, at 326 Rupees (SD = 237). Similarly, the cost of lab tests was significantly higher in Mumbai, averaging 9,970 Rupees (SD = 12,600) compared to 2,550 Rupees (SD = 3,110) in Patna.

Interestingly, the monthly cost of TB medication was lower in Mumbai (114 Rupees, SD = 372) than in Patna (188 Rupees, SD = 660), suggesting better access to subsidized medications or differences in drug pricing between the cities. More details on perspectives on the COVID-19 pandemic, how patients utilized the health care during the COVID-19 pandemic as well as cost information are provided in the supporting information (Tables C-E in S1 Text).

### Individual Care-seeking Journeys During COVID-19

In Patna, initial care-seeking predominantly began at private outpatient clinics (31%), community pharmacies (28%), and recruitment facilities (29%). Patients often continued to visit private outpatient clinics or shifted between informal providers, such as pharmacies, reflecting a fragmented journey with multiple touchpoints that could delay diagnosis and treatment. Public hospitals remained a minor player, accounting for only 9.5% of visits (Fig 3). In Mumbai, the care-seeking pathways were more streamlined, with 83% of patients initially choosing private outpatient clinics and 11% attending private hospitals or nursing homes. Subsequent visits often involved the same private providers.

In Patna, the initial healthcare encounter for TB symptoms predominantly occurred at private outpatient clinics (58%) and community pharmacies or chemists (28%), with only 11% seeking care in public hospitals. Similarly, in Mumbai, the majority of initial encounters were at private outpatient clinics (82%), followed by private hospitals (14%). The site of TB diagnosis showed a similar reliance on private facilities in both cities. In Patna, 86% of TB diagnoses were made at private outpatient clinics, whereas in Mumbai, private hospitals accounted for 55% of diagnoses, followed by private outpatient clinics at 44%.

**Fig 3.**
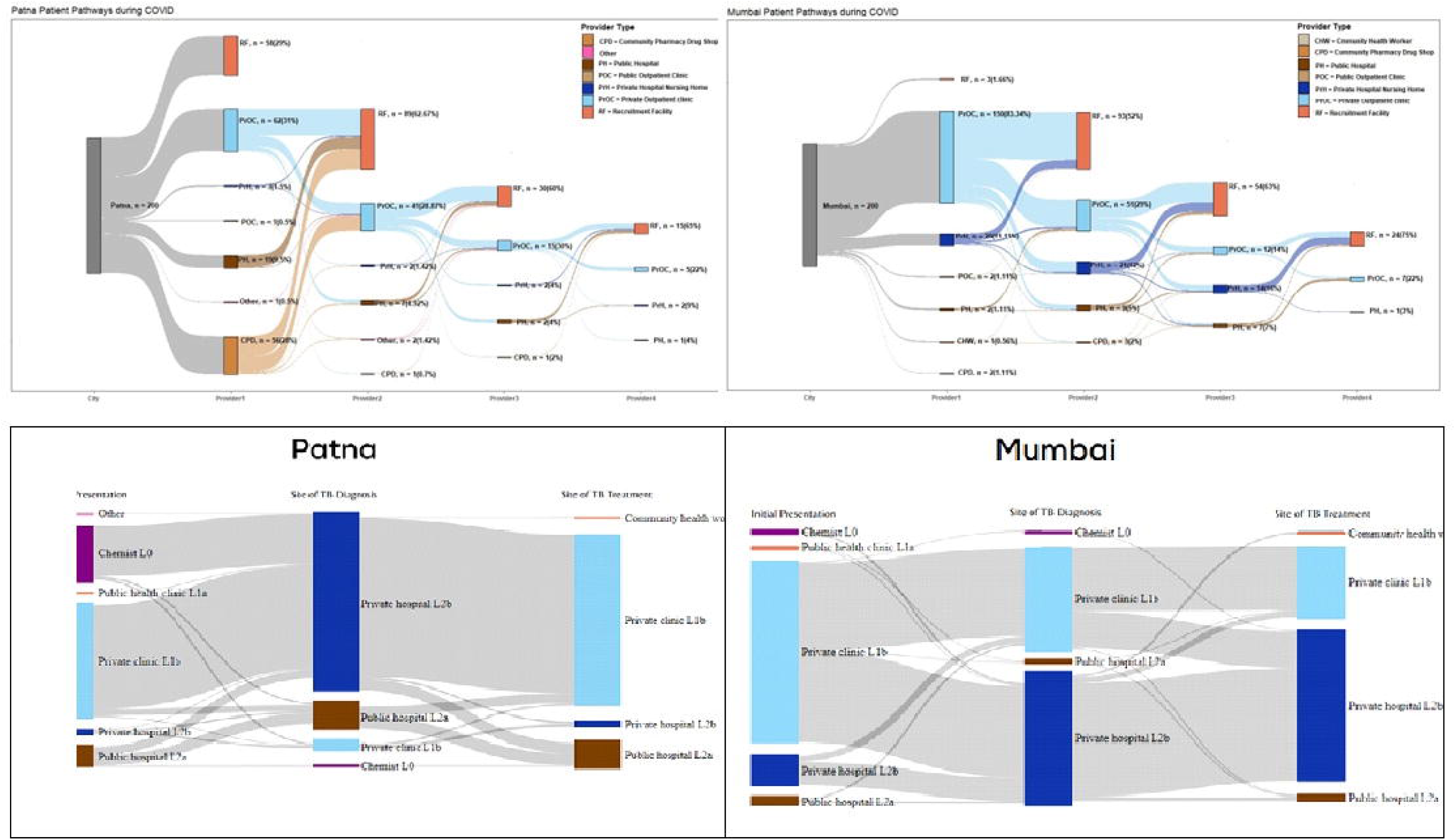
Sankey Diagrams (A - D) Showing Care-Seeking Journeys of TB Patients in Patna and Mumbai During the COVID-19.

For the initiation of TB treatment, Patna showed a strong preference for private outpatient clinics (85%), with fewer patients being treated at public hospitals (12%). In contrast, Mumbai had a higher reliance on private hospitals as the predominant site for treatment initiation (66%), indicating a greater reliance on these facilities compared to Patna. More details on patient journey and diagnostic test are provided in the supporting information (Table B in S1 Text).

### Missed Opportunities for TB Diagnosis

Fig 4 illustrates the number of healthcare visits that were missed opportunities for a confirmed TB diagnosis in Patna and Mumbai. By the second or third encounter, 75% of patients in both cities had received a diagnosis of TB. However, private outpatient clinics were associated with a higher rate of missed diagnoses, contributing to 15% of missed diagnostic opportunities. In Patna, the frequency of missed diagnoses was slightly higher, with patients requiring an average of 3 visits before diagnosis. By the third visit, 28% of patients remained undiagnosed, a higher rate than in Mumbai. Public hospitals played a smaller role in both cities, representing only 2.5% of diagnostic opportunities in Mumbai and 9.5% in Patna.

**Fig 4.**
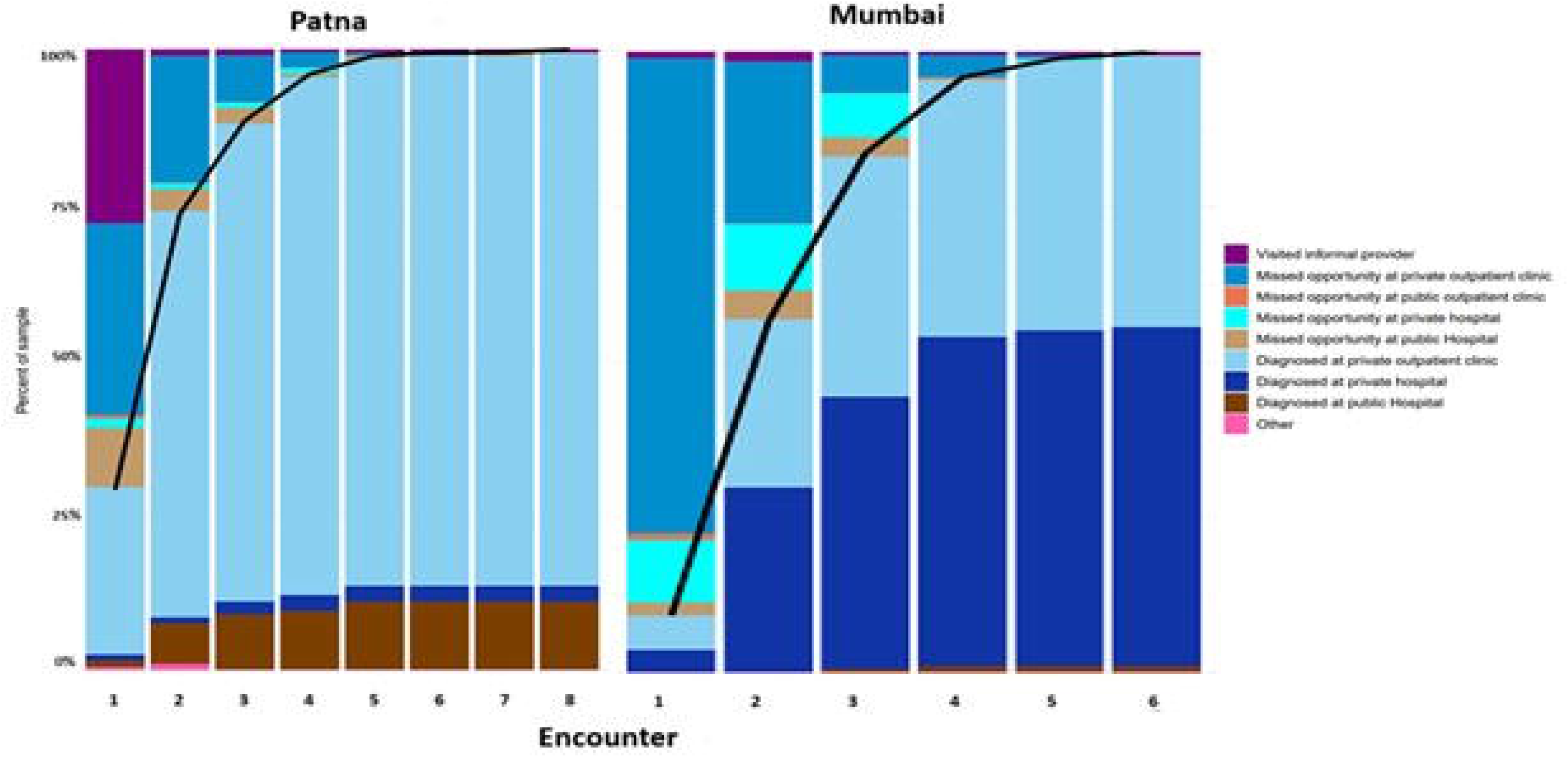
Bar charts showing sequential encounters in Patna and Mumbai during the COVID-19 pandemic.

### TB Diagnostic Testing Across Healthcare Providers

Fig 5 provides an overview of TB diagnostic testing patterns among various healthcare providers in Patna and Mumbai during the COVID-19 pandemic. In Mumbai, a higher proportion of providers recommended diagnostic tests at initial encounters compared to Patna, with 60% of first providers suggested lab tests versus 32% in Patna. The types of tests varied significantly; sputum tests were initially recommended more frequently in Patna (50%) but were less common in Mumbai at first (15%), which also decreased in later visits, reaching 0% by the fourth provider. In contrast, chest X-rays were consistently recommended in Mumbai, with 13% of first providers and 63.35% of second providers suggesting this diagnostic tool.

**Fig 5.**
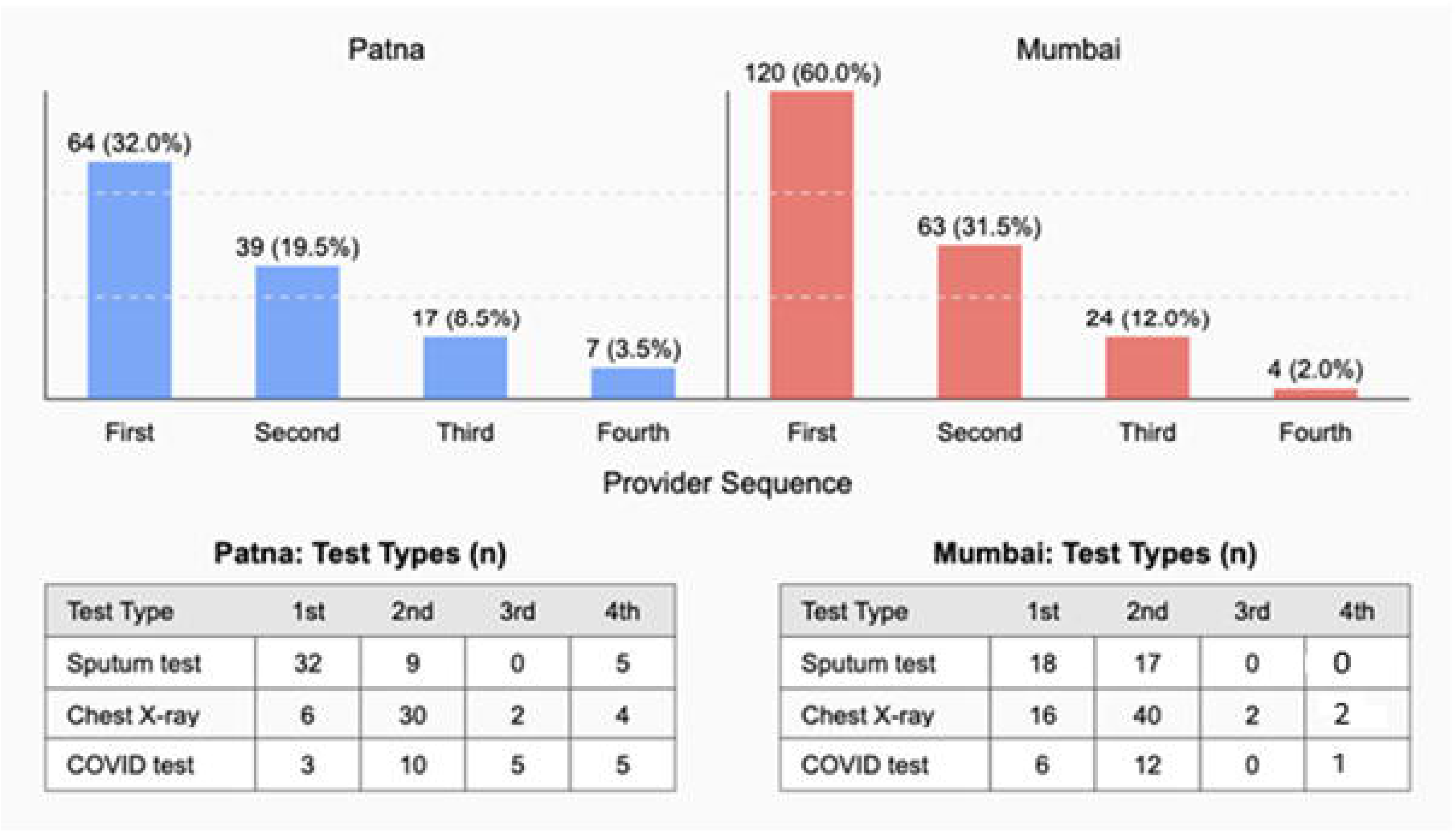
TB Diagnostic Patterns across Sequential Healthcare Providers.

Patients in Patna reported fewer difficulties in accessing recommended tests, with over 90% able to complete the tests without major barriers. In Mumbai, patients encountered more challenges over successive provider visits, with only 55.5% of patients able to complete tests without difficulty at their first encounter, and this percentage declined in subsequent visits. Sputum samples were predominantly collected at public facilities in both cities, though 23.44% of samples in Patna were also collected in private clinics or labs during the initial encounters. The timeline for receiving sputum test results differed between the cities. In Patna, a greater proportion of patients (20.32%) received results within 1-2 days, compared to only 6% in Mumbai.

### Factors Associated with Diagnostic and Treatment Delays

Figs 6a and 6b presents the analysis of the factors associated with median change in healthcare-seeking, provider, treatment, and total delays (measured in days) during the COVID-19 pandemic in Patna and Mumbai. In Patna, for healthcare-seeking delay, males reported slightly shorter delay compared to females (adjusted median: -0.79 days; 95% CI: -2.26, 0.67; p=0.290), and individuals with inadequate financial resources showed negligible difference (adjusted median: 0.25 days; 95% CI: -1.89, 2.39; p=0.819). Provider delay was lower among males (adjusted median: -7.92 days; 95% CI: -43.68, 27.85; p=0.665) and those who sought care at public hospitals (24.58 days, 95% CI: -64.87, 15.71; p = 0.234), though these differences were not statistically significant. Treatment delays were higher among males (adjusted median: 2.23 days; 95% CI: -1.68, 6.14; p=0.266), while marital status and education level showed no notable influence. Total delay was slightly lower among males (adjusted median: 2.40 days; 95% CI: - 52.23, 57.04; p=0.931) and marginally shorter among individuals with bachelor’s degrees (adjusted median: -5.27 days; 95% CI: -74.45, 63.91; p=0.882). Details of the unadjusted and adjusted analyses are presented in the supporting information (Tables F-M in S1 Text).

**Fig 6a.**
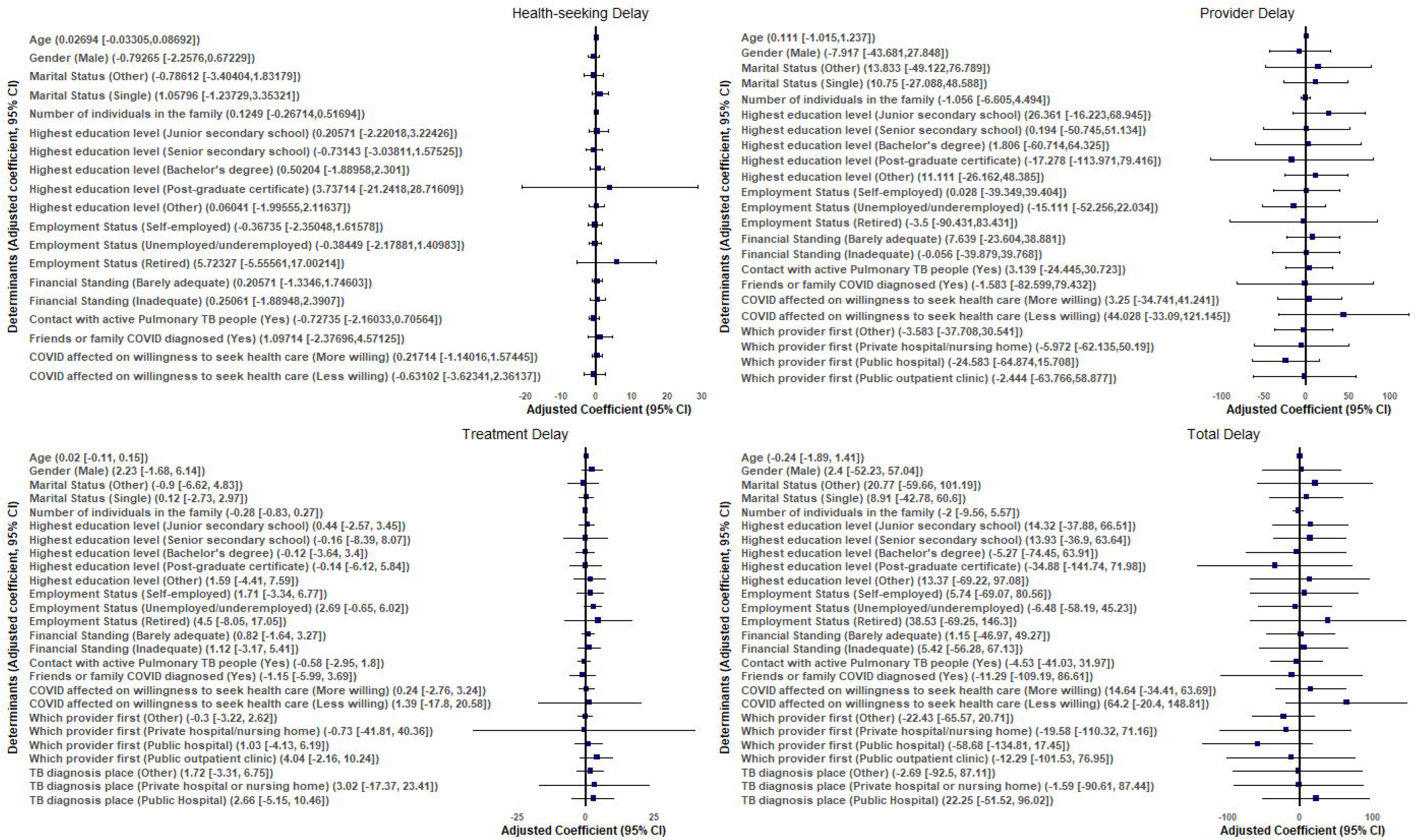
Factors associated with Diagnostic and Treatment Delays during the COVID-19 Pandemic in Patna.

**Fig 6b.**
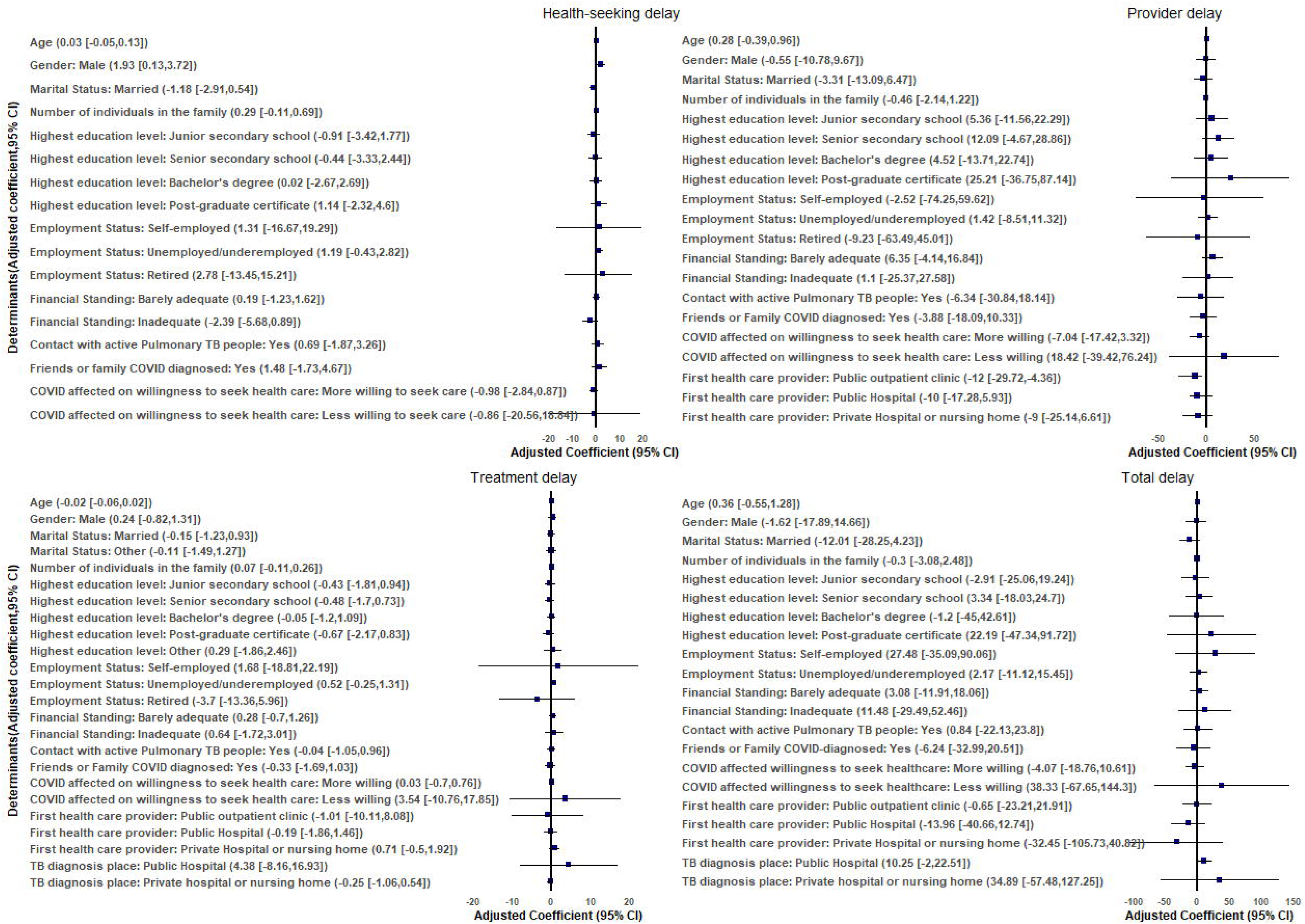
Factors associated with Diagnostic and Treatment Delays during the COVID-19 Pandemic in Mumbai.

In Mumbai, men experienced longer delays in seeking care than women (adjusted median: 1.93 days; 95% CI: 0.13, 3.72; p=0.037). Household size showed a trend toward increased delays, with a 0.29-day increase (95% CI: -0.11, 0.69; p=0.080). Health seeking delays were also higher among unemployed individuals (adjusted median: 1.19; 95% CI: -0.43,2.82; p=0.152) and those with adequate financial resources compared to those with inadequate means (adjusted median: -2.39 days; 95% CI: -5.68, 0.89; p=0.155). In the case of provider delay, being married reduced this delay compared to single individuals (adjusted median: -3.31, 95% CI: -13.09,6.47; p=0.508). Unemployed people had higher provider delays than those employed (adjusted median: 1.42; 95% CI: -8.51,11.32; p=0.782), while initial visits to public outpatient clinics were associated with a decrease in provider delay (adjusted median: -12, 95% CI: -29.72, 4.36; p=0.871).Treatment delay was marginally higher among males (adjusted median: 0.24, 95% CI: -0.82, 1.31; p=0.232), and unemployed individuals (adjusted median: 0.52, 95% CI: -0.25, 1.31; p=0.194), though these differences were not statistically significant. For total delay, gender, marital status, and the household size showed no significant impact; however, unemployed individuals faced a slightly higher delay (adjusted median: 2.17; 95% CI: -11.12,15.45; p=0.777). A greater delay was observed among those less willing to seek care (adjusted median: 38.33; 95% CI: -67.65,144.30; p=0.401).

### Factors Associated with Number of Healthcare Encounters

In Mumbai, a larger household size was associated with an increased number of healthcare interactions (adjusted coefficient: 0.08, 95% CI: 0.02, 0.14; p = 0.009). Unemployed participants had fewer encounters (adjusted coefficient: -0.38, 95% CI: -0.72, -0.05; p=0.025), while individuals with barely adequate resources had an increased number of encounters (adjusted coefficient: 0.35, 95% CI: 0.02, 0.67; p=0.038) (Table 4).

A greater distance to both public and private healthcare was associated with lower numbers of encounters (adjusted coefficient: -0.28, 95% CI: -0.68, 0.12; p = 0.166) and (adjusted coefficient: -0.07, 95% CI: -0.98, 0.84; p = 0.882), respectively. Weekly internet use for health information was associated with a non-significant increase in encounters (adjusted coefficient: 1.23, 95% CI: -0.27, 2.73; p = 0.107). While contact with active pulmonary TB patients, having friends or family diagnosed with COVID-19, and varying willingness to seek healthcare due to COVID-19 were not significant, those reporting a lower willingness to seek care showed a slight, significant increase (adjusted coefficient: 0.89, 95% CI: 0.15, 1.64; p = 0.019) (Table 4).

**Table 4.**
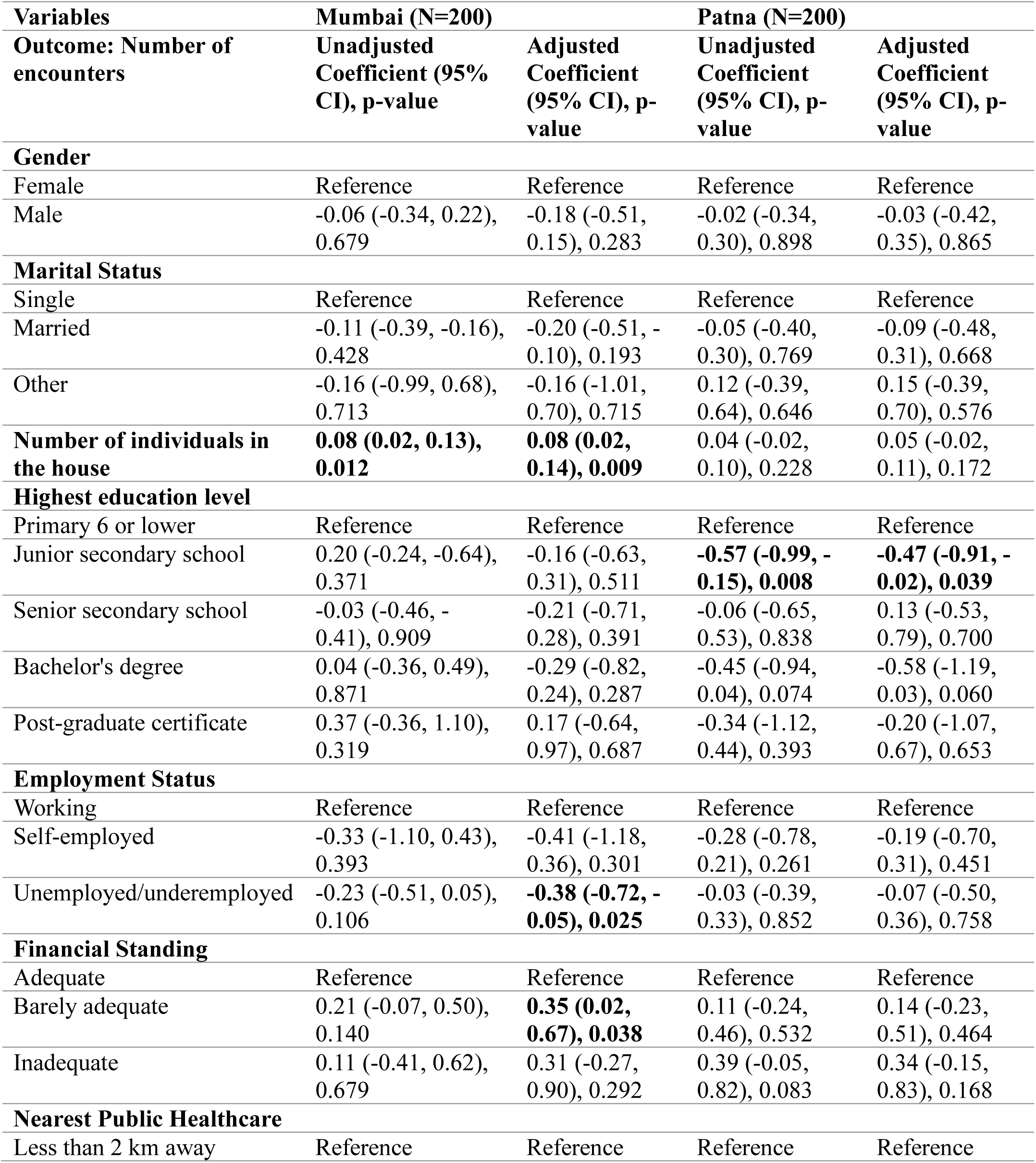
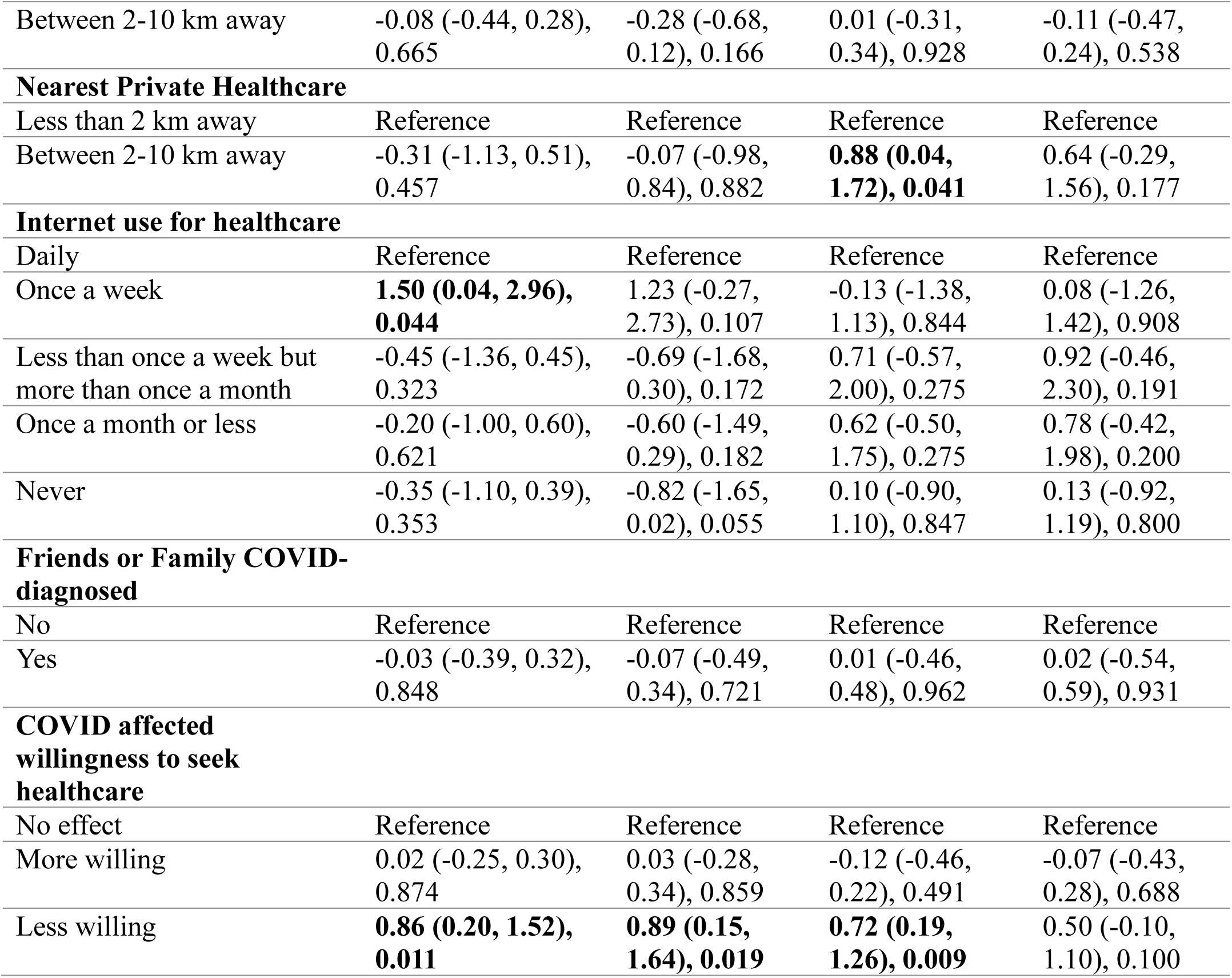
Linear regression: factors associated with the number of encounters in Mumbai and Patna.

In Patna, gender and marital status were not significantly associated with healthcare encounters. Participants with junior school education or those studying for a bachelor’s degree reported fewer encounters compared to those with only primary education (adjusted coefficient: -0.47, 95% CI: - 0.91, -0.02; p = 0.039) and those with secondary education (adjusted coefficient: -0.58, 95% CI: - 1.19, 0.03; p = 0.060), respectively. Employment status was not a strong predictor, while those reporting inadequate resources showed a non-significant trend toward a greater number of encounters (adjusted coefficient: 0.34, 95% CI: -0.15, 0.83; p = 0.168).

The distance to the nearest public healthcare facility had no significant impact on the number of encounters. In contrast, a greater distance to private healthcare facilities was associated with a potential increase in encounters (adjusted coefficient: 0.64, 95% CI: -0.29, 1.56; p = 0.177). The frequency of internet use for health information and COVID-19-related factors, such as contact with active TB patients or having family or friends diagnosed with COVID-19, was not significantly associated with the number of healthcare encounters. However, those who were less willing to seek healthcare due to COVID-19 showed a trend toward an increased number of encounters (adjusted coefficient: 0.50, 95% CI: -0.10, 1.10; p = 0.100).

## DISCUSSION

The convergence of the TB epidemic and the COVID-19 pandemic posed dual challenges for healthcare systems, especially in resource-limited settings like India, creating unprecedented strain. Our study examined how TB patient journeys, healthcare access and associated costs in Mumbai and Patna, were affected by the COVID-19 pandemic. The results offer critical insights into care delays, patient behaviors, healthcare utilization patterns, and financial challenges, underscoring the urgent need for targeted public health measures to mitigate these adverse effects and build more resilient health systems for the future.

### Delays in Care and Factors Influencing Healthcare-Seeking Behavior

Healthcare-seeking behaviors during crises are significantly influenced by demographic and socioeconomic factors, as documented across multiple settings [39]. Gender-based disparities in care access during the pandemic have been observed globally, with several studies noting shifts in typical patterns [40]. In Mumbai, men experienced longer delays compared to women, reflecting gender disparities consistent with cultural norms and previous research [39]. Larger household sizes in Mumbai were associated with increased healthcare interactions, possibly due to heightened vigilance among families at risk. Research from other high-burden settings similarly suggests that the pandemic altered usual care-seeking behaviors, potentially exacerbating pre-existing inequities in healthcare access [41].

Unemployment contributed to longer delays in both cities, suggesting that financial insecurity during the pandemic hampered timely access to care. The complex relationship between economic resources and healthcare utilization during crises has been extensively documented by multiple researchers, who have consistently found that individuals with extremely limited resources often forgo care entirely [42]. In contrast, those with constrained but not absent resources may make extraordinary efforts to access healthcare despite significant barriers. Interestingly, individuals with limited but adequate resources sought more healthcare encounters, possibly driven by a heightened perceived risk despite economic constraints. This paradoxical trend underscores the importance of financial security in healthcare utilization [43,44].

### Impact of Healthcare Provider Type on Diagnostic and Treatment Delays

The role of private healthcare providers in care delays during emergencies has been increasingly recognized in health systems research [45]. Studies from several South Asian contexts have documented how reliance on private providers shapes care pathways, particularly in urban settings [32]. This phenomenon is especially pronounced in India, where reliance on private healthcare reflects longstanding perceptions about quality and accessibility but creates substantial financial vulnerability during crises [32].

The predominance of private healthcare providers played a significant role in care delays. In Patna, initial visits to public hospitals were associated with shorter provider delays. In contrast, Mumbai’s reliance on private hospitals resulted in higher consultation fees and potential delays due to overwhelmed capacity. This reliance on private healthcare, especially in Mumbai, reflects perceptions of better quality and proximity but has led to substantial patient financial burdens [5], this reliance on private healthcare during the pandemic resulted in substantial financial burdens, with patients facing high out-of-pocket expenses [46,47]. The disparities in diagnostic costs between the cities further compounded challenges, particularly for lower-income households disproportionately affected by increased healthcare expenses [8].

### Economic Impact of COVID-19 on TB Patients

The pandemic’s economic impact on healthcare costs has been widely documented. Research from multiple settings has demonstrated how health emergencies can exacerbate the economic burden of chronic disease care [47], with particularly severe effects on socioeconomically vulnerable populations [8]. Studies from Nigeria and Peru have similarly documented how financial barriers during the COVID-19 pandemic delayed healthcare access and treatment adherence for TB patients [43], suggesting this is a widespread phenomenon that requires policy attention.

The economic burden of TB care intensified during the COVID-19 pandemic, particularly in Mumbai, where 85.5% of participants reported higher healthcare costs driven by elevated consultation and diagnostic fees. These financial strains likely delayed healthcare access and treatment adherence, echoing trends seen in other settings during the pandemic [43] Policies to reduce out-of-pocket expenses, such as expanding public healthcare subsidies or providing financial aid for diagnostics, could mitigate these barriers.

### Policy Implications

The COVID-19 pandemic offers important lessons about health system vulnerabilities worldwide. Global health experts have noted how the pandemic exposed critical weaknesses in essential service delivery that extend beyond the acute crisis period [48]. The documented disruptions to TB care during COVID-19 mirror patterns seen during previous health emergencies, suggesting the need for fundamental strengthening of health service delivery models [49].

Building more resilient health systems requires addressing several key dimensions. Research on health system strengthening emphasizes the importance of integrating public and private healthcare sectors to ensure coordinated responses during crises [32] and implementing financial protection mechanisms that prevent catastrophic expenditures [50]. The WHO and the Stop TB Partnership have highlighted the importance of developing emergency preparedness plans specifically for maintaining essential services, which can protect vulnerable populations during future crises [51]. Evidence from multiple settings suggests that addressing socioeconomic and gender-based barriers through targeted interventions is crucial for building equitable and resilient systems capable of withstanding future shocks [49].

Our findings underscore the need for policy interventions to strengthen TB services and prepare health systems for future disruptions. Strengthening public healthcare systems, particularly in high-burden urban areas like Mumbai, can reduce reliance on costly private care and improve access to affordable health services. Integration between the public and private sectors and investments in public healthcare infrastructure can foster a more resilient system capable of managing concurrent health emergencies [52] Reducing out-of-pocket expenses, such as subsidizing medications and diagnostic tests and enhancing public health insurance, would financially relieve vulnerable populations. Addressing gender-specific barriers, as evidenced by longer delays among men in Mumbai, is another priority. Public health campaigns tailored to cultural norms can promote equitable healthcare access and improve diagnostic timeliness.

Investment in digital health solutions, including telemedicine and mobile health (mHealth) tools, has been increasingly recognized as a strategy to bridge gaps in healthcare access [53,54]. By leveraging these technologies for patient education and remote monitoring, health systems can maintain the continuity of TB care even when in-person services are disrupted [53,54]. By promoting digital literacy and expanding telehealth services, policymakers can enhance healthcare accessibility, reduce delays, and minimize costs during emergencies. Leveraging these tools for patient education and monitoring will ensure continuity of care even in times of crisis.

### Limitations

This study’s strengths include its comprehensive analysis of patient pathways, the economic impact of healthcare costs, and factors influencing healthcare interactions during the COVID-19 pandemic. Using both quantitative and qualitative data enabled a nuanced understanding of the challenges faced by TB patients in Mumbai and Patna. However, there are limitations to consider. The cross-sectional design limits the ability to establish causality, and the self-reported nature of some variables may introduce recall bias. Additionally, the study focused on two urban centers, which may not to be generalizable to rural areas where healthcare access and economic conditions differ significantly. The potential for recall bias in self-reported healthcare-seeking behaviors has been acknowledged in similar studies [55] and cross-sectional designs have inherent limitations for establishing causal relationships. Additionally, the generalizability of urban-focused research to rural areas requires careful consideration, though the patterns observed often highlight important systemic issues relevant to broader TB control efforts.

## CONCLUSION

Our study underscores the complexities and challenges TB patients faced in India during the COVID-19 pandemic, highlighting the critical need for adaptive, patient-centered healthcare systems. The COVID-19 pandemic revealed vulnerabilities in TB care journeys in urban India that provide important lessons for health system resilience beyond the immediate crisis. Strengthening the integration between public and private healthcare sectors, reducing financial barriers, leveraging technology for healthcare delivery, and promoting equitable access to care is imperative. By positioning COVID-19’s impact as a case study of health system disruption, we demonstrate how diagnostic, and treatment delays reflect structural weaknesses that persist beyond the acute crisis period. This study contributes valuable insights into the complexities of managing TB during health system disruptions, offering directions for future research and policy-making that mitigate the impact of concurrent health crises on vulnerable populations and strengthen system-wide preparedness for maintaining essential services during future emergencies.

## Data Availability

All data produced in the present study are available upon reasonable request to the authors.

## Acknowledgments

We are grateful to Ranen Das, Charu Nanda and the ISERDD field team for their valuable work in data collection. We also thank Caroline Vadnais, from the Research Institute of the McGill University Health Centre, for outstanding administrative support.

## Notes

### Competing Interest Statement

The authors have declared no competing interest.

### Funding Statement

This study was funded by the Bill and Melinda Gates Foundation (Grant #: INV-022420; PIs: Madhukar Pai, Jishnu Das). The funders had no role in the study design, data collection and analysis, the decision to publish, or the preparation of the manuscript.

### Author Declarations

McGill University Research Ethics Board (Covid BMGF / 2021-7197), the Georgetown-Medstar IRB (STUDY00003422), and the Institute for Social and Economic Research on Development and Democracy (ISERDD) gave ethical approval for this work.

