## Supplemental materials for "Delays in tuberculosis diagnosis and treatment in India: A patient journey analysis from Mumbai and Patna"

**S1 Text**

### **Table A. Patient Journey Survey (Questionnaire)**

| **#** | **Question** | **Options** | **Skip pattern** |
| --- | --- | --- | --- |
| **Section 1: Profile of the individual** | | | |
| 1. | Date of birth | (DD/MM/YYYY) |  |
| 2. | What is your gender? | a. Male b. Female c. Other d. Prefer not to say |  |
| 3. | What is your current marital/partner status? | a. Single b. Married c. Divorced d. Separated e. Widowed f. Prefer not to say |  |
| 4. | What type of dwelling do you currently live in? | a. Own house/apartment b. Rented house/apartment c. Informal housing (huts, d. Other, specify: ______ |  |
| 5. | Do you (or your spouse/partner) own or rent your dwelling? | a. Own outright b. Own on mortgage c. Rent from local authority/housing association d. Rent from private landlord e. Other, specify: ______ |  |
| 6. | How many individuals (relationship, age, and gender) live with you? | #_________ **For each person that lives with you specify their relationship to you, their age, and their gender:** Relationship \| Age \| Gender |  |
| 7. | What is the highest level of education that you have obtained? | a. Primary 6 or lower b. Junior secondary school c. Senior secondary school d. Trade certification or diploma from a vocational school e. OND/HND f. Bachelor’s degree g. Post-graduate certificate or degree above bachelor’s degree h. Other, specify: ______ |  |
| 8. | What is your current employment status? | a. Working b. Self-employed c. Unemployed/underemployed d. Retired | If c, skip to 10.  If d, skip to 12. |
| 9. | Which of the following describes your working schedule? | a. Daytime shift b. Evening shift c. Night shift d. Rotating shift e. Seasonal, on-call, no pre-arranged schedules f. Other, specify: ______ | After this question, skip to 12. |
| 10. | If not working, how long have you been unemployed? | a. Unemployed as of this month b. 1-2 months c. 2-6 months d. Over 6 months e. Never been employed |  |
| 11. | What would best describe the reason for not working? | a. Unable to work because of sickness or disability b. Looking after family c. Student d. Doing unpaid work e. Laid off due to COVID-19 pandemic f. Laid off due to some other reason than COVID-19 pandemic g. Other, specify: ______ |  |
| 12. | How would you describe your **current** financial standing? | a. Adequate b. Barely adequate c. Inadequate |  |
| 13. | How would you describe your financial standing **before the start of the COVID-19 Pandemic in March 2020**?  **Read answer options** | a. Adequate (my family and I can meet our needs every month) b. Barely adequate (e.g., some months my family and I can meet our needs, others we cannot) c. Inadequate (e.g., most months my family and I cannot meet our needs) |  |
| 14. | Which of these phrases best describes how you (and your spouse/partner) are getting along financially now (during the COVID-19 pandemic)? | a. Manage very well b. Manage quite well c. Get by alright d. Don’t manage very well e. Have some financial difficulties f. Have severe financial difficulties |  |
| 15. | How far away from your home is the nearest public health centre? | a. Less than 2 km away b. Between 2-10 km away c. Over 10km d. Don’t know/not sure |  |
| 16. | How far away from your home is the nearest private clinic, nursing home, or hospital? | a. Less than 2 km away b. Between 2-10 km away c. Over 10km a. Don’t know/not sure |  |
| 17. | How often do you use the internet for accessing healthcare? i.e. investigating symptoms, consultations on what medications to take, learning about prognosis of past diseases | a. Daily b. More than once a week c. Once a week d. Less than once a week but more than once a month e. Once a month or less f. Never g. Other, specify: _______ |  |
| 18. | Do you have a history of contact with people with active Pulmonary TB? | a. Yes b. No |  |
| **Section 2: Perspectives on the COVID-19 pandemic** | | | |
| 19. | At any point between March 2020 and now, have there been any COVID-19 lockdowns or restrictions in place around where you live/your neighbourhood? | a. Yes b. No c. Don’t know |  |
| 20. | Which of the following COVID-19 related restrictions or protocols **are currently in place** you live/in your neighbourhood?  **Read answer options and check** **all that apply** | a. Movement completely restricted, full lockdown (e.g., all stores, restaurants, bars, churches, and mosques closed) b. Restaurants and bars are open with limited capacity c. Markets and stores open with limited capacity d. Religious ceremonies and services at churches and mosques at limited capacity e. Movement partially restricted (e.g., curfew, daily walks/exercise permitted) f. Masks mandatory in public spaces g. Social distancing mandated h. Large gatherings are restricted i. Other, specify:________ |  |
| 21. | Which of the following COVID-19 related restrictions or protocols **have ever been in place** you live/in your neighbourhood?  **Read answer options and check** **all that apply** | a. Movement completely restricted, full lockdown (e.g., all stores, restaurants, bars, churches, and mosques closed) b. Restaurants and bars are open with limited capacity c. Markets and stores open with limited capacity d. Religious ceremonies and services at churches and mosques at limited capacity e. Movement partially restricted (e.g., curfew, daily walks/exercise permitted) f. Masks mandatory in public spaces g. Social distancing mandated h. Large gatherings are restricted i. Other, specify: ______ |  |
| 22. | Which statement best describes your overall experience accessing healthcare during the COVID-19 pandemic (compared your access before the pandemic)?  **Read answer options and check all that apply** | a. It is easier to access healthcare b. It is more difficult to access healthcare c. Access to healthcare stayed the same |  |
| 23. | Select the statement(s) that reflect how COVID-related lockdowns or restrictions have affected your access to healthcare since the pandemic started in March 2020.  **Read answer options and check** **all that apply** | a. I was unable to reach a doctor because facilities were closed due to the COVID-19 pandemic b. I was unable to receive my medications because drug stores were closed due to the COVID-19 pandemic c. I had to seek care from a different facility than my typical/preferred facility d. Waiting times at facilities were longer than normal/expected e. I was unable to leave the house to seek care because of movement/transport restrictions f. Other, specify:______________ |  |
| 24. | How frequently do you go outside your house/apartment/flat? | a. Daily b. Less than daily but more than once a week c. Once a week d. Less than once a week |  |
| 25. | Which of the following are the reason(s) you go outside of your house/apartment/flat?  **Read answer options and check** **all that apply** | a. Go to the market or other errands b. Seek healthcare or medicine c. Drop off/pick-up children from school/nursery d. Work e. School f. Exercising g. Socializing with family, neighbours, or friends h. Attend religious gathering at a church, mosque, etc. i. Other, specify:__________ |  |
| 26. | Have **you or any of your close friends and family** been diagnosed with COVID-19? | a. Yes b. No | If b, skip to 27. |
| 27. | Choose the statement(s) which characterises **your personal or immediate family experience** with COVID-19.  **Read answer options and check** **all that apply** | a. Diagnosed with COVID-19 but experienced mild/moderate symptoms b. Very sick with COVID-19 for a short period (Two weeks or less) c. Very sick with COVID 19 for a long period (More than two weeks) d. Hospitalized with COVID 19 e. Death due to COVID 19 (in case of a loved one) f. Other, specify |  |
| 28. | Has COVID and/or current/past COVID-related restrictions affected *your willingness* to seek healthcare? | a. Yes, I am more willing to seek care b. Yes, I am less willing to seek care c. No effect d. Don’t know/not sure | If a, skip to 29.  If b, skip to 28.  If c or d, skip to 30 |
| 29. | Select the statement(s) that describe how COVID has *decreased* your willingness to seek health care.  **Read answer options and check** **all that apply** | a. I am afraid of visiting health facilities because of a fear of exposure to COVID-19 infection b. I am afraid of visiting health facilities because I don’t want to be tested or diagnosed with COVID-19 infection c. I am less able to/can no longer afford to seek care because of lost income in the past year d. Other, specify:__________ |  |
| 30. | Select the statement(s) that describe how COVID has *increased* your willingness to seek health care.  **Read answer options and check** **all that apply** | e. I am more aware that early healthcare seeking is important f. I have to get tested regularly for work/school g. I am worried about infecting my family and friends therefore I seek healthcare more often h. I am worried about getting infected from people I come in contact with therefore I seek healthcare more often i. I want to see my family and therefore as a precaution I get tested and seek extra care j. My underlying/chronic condition makes me more willing to seek care during the COVID-19 pandemic   Other, specify |  |
| **Section 3: Health Care Utilization during the COVID-19 pandemic** | | | |
| 31. | In general, how would you describe your overall health (including mental health)?  **Read answer options** | a. Excellent b. Very good c. Good d. Fair e. Poor |  |
| 32. | Compared with before March 01, 2020, how would you describe your health now?  **Read answer options** | a. Better b. Worse c. About the same |  |
| 33. | Including this facility, what are all the different types of health providers you have had contact with about your physical or mental health since the beginning of the COVID-19 pandemic?  **Read answer options and check all that apply.**  **Remind patient they should count the current facility, in addition to any others consulted since March 2020.** | a. Community health worker/CHEW b. Community pharmacy, chemist, drug shop, patent medicine vendor (PPMV) c. Private laboratory d. Public outpatient clinic (e.g., PHC /Health Post/Dispensary) e. Public Hospital f. Private outpatient clinic, health centre, or dispensary g. Private hospital or nursing home h. Emergency room in a public facility i. Emergency room in a private facility j. Traditional healers k. Other, specify: ______ |  |
| 34. | When you get sick and you decide to seek treatment, where do you typically go to seek help first?  **Read answer options** | a. Community health worker/CHEW b. Community pharmacy, chemist, drug shop, patent medicine vendor (PPMV) c. Private laboratory d. Public outpatient clinic (e.g., PHC /Health Post/Dispensary) e. Public Hospital f. Private outpatient clinic, health centre, or dispensary g. Private hospital or nursing home h. Emergency room in a public facility i. Emergency room in a private facility j. Traditional healers k. Other, specify: ______ |  |
| 35. | How has the health facility you go to most frequently been affected by the COVID-19 pandemic?  **Read answer options and check all that apply** | a. Closed b. Reduced hours c. Reduced services d. Screening for COVID-19 e. Increased cost f. Decreased cost g. Infection control measures (spacing (as in physicial distancing), mandatory mask-wearing, hand washing, hand sanitizing stations, etc.) h. No change/has not been affected i. Other, specify: _______ |  |
| 36. | Since March 01, 2020, was there ever a time when you felt that you needed health care but didn’t receive it? | a. Yes b. No c. Can’t remember | If b or c, skip to 37 |
| 37. | Thinking of the most recent time, why did you not receive it?  **Read answer options and check all that apply** | a. Healthcare was not available because practice was closed due to the COVID-19 pandemic b. Healthcare was not available because practice was closed, but not due to COVID-19 c. Waiting time was too long d. Could not afford e. Too busy with work and other responsibilities f. Decided not to seek care due to fear of exposure to COVID-19 pandemic g. Other, specify: _____ |  |
| 38. | Since the start of the COVID pandemic in March 2020, have you had any problems obtaining any non-TB-related medications you needed? | a. Yes b. No c. Can’t remember | If b or c, skip to 39 |
| 39. | Which of the following best describe the reasons you have had trouble obtaining any non-TB-related medicines?  **Read answer options and check all that apply** | a. Pharmacy/provider was closed  b. Pharmacy/provider was understocked c. I had problems getting to the pharmacy d. I could not afford the medicine e. I don’t have health insurance f. Other problem, specify: _______ |  |
| 40. | We would like to know how frequently you have used telemedicine. Telemedicine includes using phone calls, video calls, WhatsApp, or online “virtual visits” to consult with a health care provider.  **Before the start of the COVID-pandemic**, how frequently were you using telemedicine to consult with a health care professional? | a. Did not use telemedicine b. Daily c. More than once a week d. Once a week e. Less than once a week but more than once a month f. Once a month or less g. Other, specify:__________ |  |
| 41. | **Since the start of COVID-pandemic** in March 2020, how frequently have you been using telemedicine to consult with a health care professional? | a. Did not use telemedicine b. Daily c. More than once a week d. Once a week e. Less than once a week but more than once a month f. Once a month or less g. Other, specify: _______ |  |
| **Section 4: TB Diagnosis and treatment initiation** | | | |
| 42. | Please confirm, have you been diagnosed with TB?  (e.g. you have received test results that have said you are positive for TB) | a. Yes b. No c. Not sure |  |
| 43. | Which of these statements best describes the stage of TB care you are in right now?  **Read answer options** | a. My doctor told me I have TB but I have not been given any medicines yet b. I started taking TB medication, but I stopped before finishing all my medication c. I am currently taking TB medication and have been taking my medicines up to today d. I finished taking all my TB medication e. Not sure f. Other, specify: _________ |  |
| **Section 5: onset of symptoms and care seeking pathway** | | | |
| 44. | Now I’d like to ask you some questions about the illness that led you to seek care at this facility.  Thinking back to when you first started feeling ill, which of the following symptoms prompted you to seek help from a health provider?  **Read answer options and check** **all that apply** | a. Cough b. Chest congestion c. Head congestion d. Sore throat e. Rashes or allergies f. Trouble breathing/wheezing g. Weight loss h. Night sweats i. Fever j. Lack of appetite k. Other, specify ____________ |  |
| 45. | Thank you for telling me about your symptoms.  Approximately how much time passed from when you first noticed these symptoms and when you decided you needed to seek help from a health care provider? | a. 2 days or less b. 3-7 days (up to 1 week) c. More than 1 week but less than 1 month d. 1-2 months e. 2-3 months a. Over 3 months |  |
| 46. | ***Before coming to this facility***, did you seek care from any other healthcare providers? | b. Yes c. No f. |  |
| 47. | How many other providers did you seek care from **before coming to this facility**? | #_________________ |  |
| 48. | Now I am going to ask you about each of the providers you visited *before coming to this facility.*  After you decided to seek care for your symptoms, which type of provider did you go to **[first]**?  **Read answer options** | a. Community health worker/CHEW b. Community pharmacy, chemist, drug shop, patent medicine vendor (PPMV) c. Public outpatient clinic (e.g., PHC /Health Post/Dispensary) d. Public Hospital e. Private outpatient clinic, health centre, or dispensary f. Private hospital or nursing home g. Emergency room in a public facility h. Emergency room in a private facility i. Traditional healers   Other, specify: ______ |  |
| 49. | Did the [**first provider**] tell you that you needed a lab test for your symptoms? | 1. Yes 2. No   Don’t know/can’t remember |  |
| 50. | What type of lab test(s) or diagnostic(s) did the [**first provider**] recommend?  **Read answer options and check** **all that apply** | a. Sputum test (e.g., testing mucus/phlegm using smear microscopy, AFB, GenXpert, MCS) b. Chest X-ray c. COVID-test (nasal/oral swab) d. Blood test e. Unknown f. Other, specify: ______ |  |
| 51. | Were you able to get these tests done? | a. Yes, with no difficulties b. Yes, with difficulties c. No, lab was closed d. No, I couldn’t produce sputum e. No, I didn’t have time f. Don’t know/don’t remember g. Other, specify |  |
| 52. | In the case of a sputum test, where was sputum collected? | a. At a private clinic/testing centre/ laboratory b. At a public facility/testing centre/laboratory c. At a mobile testing centre/camps Sputum collected at home d. Sputum test recommended, but patient did not follow through with the test. e. Other, specify: ______ |  |
| 53. | How long did it take to receive the results of your sputum test from the [**first provider**]? | a. Same day, or 1-2 days b. 3-7 days (up to 1 week) c. More than 1 week but less than 1 month d. 1-2 months e. 2-3 months f. Over 3 months g. Never received my result |  |
| 54. | How did the [**first provider**] communicate the sputum test results of the test with you?  **Read answer options** | a. In person at the health facility b. They called me c. They sent me a text message d. They sent me an e-mail e. Other, specify:__________ |  |
| 55. | How much time passed between when you saw the [**first provider**] and when you decided to go to the next provider? | a. 1 or 2 days b. 3-7 days (up to 1 week) c. More than 1 week but less than 1 month d. 1-2 months e. 2-3 months   Over 3 months |  |
| Repeat question sequence Q47-54 for each additional health care provider the patient visited prior to current facility, based on Q46 (e.g., if visited 3 providers before the current facility, repeat Q35-48 three times) | | | |
| 56. | Where were you **diagnosed** with TB? | a. Community health worker/CHEW b. Community pharmacy, chemist, drug shop, patent medicine vendor (PPMV) c. Public outpatient clinic (e.g., PHC /Health Post/Dispensary) d. Public Hospital e. Private outpatient clinic, health centre, or dispensary f. Private hospital or nursing home g. Emergency room in a public facility h. Emergency room in a private facility  j. Traditional healers   Other, specify: ______ |  |
| 57. | Is the facility where you were diagnosed with TB the same as the first health facility where you went to after you noticed your symptoms? | a. Yes b. No |  |
| 58. | Where do you receive TB treatment? | a. Community health worker/CHEW b. Community pharmacy, chemist, drug shop, patent medicine vendor (PPMV) c. Public outpatient clinic (e.g., PHC /Health Post/Dispensary) d. Public Hospital e. Private outpatient clinic, health centre, or dispensary f. Private hospital or nursing home g. Emergency room in a public facility h. Emergency room in a private facility  k. Traditional healers   Other, specify: ______ |  |
| 59. | Is the place of TB treatment the same as the facility where you were diagnosed? | a. Yes b. No |  |
| 60. | Now I’d like to ask you some questions about your experience *in THIS facility*.  Has your *current provider* told you that you needed a lab test or diagnostic for your symptoms? | a. Yes b. No   Don’t know/can’t remember |  |
| 61. | What type of lab test or diagnostic did your *current provider* recommend?  **Read answer options and check** **all that apply** | a. Sputum test (e.g. smear microscopy, AFB, GenXpert, MCS) b. Chest X-ray c. COVID-test d. HIV test e. Malaria test f. Typhoid test g. Blood test h. Unknown i. Other, specify: ______ |  |
| 62. | Were you able to get these tests done? | a. Yes, with no difficulties b. Yes, with difficulties c. No, lab was closed d. No, I couldn’t produce sputum e. No, I didn’t have time f. Don’t know/don’t remember g. Other, specify | If e, go to next question. If not, skip to 58. |
| 63. | In the case of a sputum test, where was sputum collected? | a. Here in this facility b. At a public facility/testing centre/laboratory c. At a mobile testing centre/camps d. Sputum collected at home e. Sputum test recommended, but patient did not follow through with the test   Other, specify: ______ |  |
| 64. | How long did it take to receive the results of your sputum test from your ***current provider***? | a. Same day, or up to 2 days b. 3-7 days (up to 1 week) c. More than 1 week but less than 1 month d. 1-2 months e. 2-3 months f. Over 3 months g. N/A – patient did not do sputum test with this provider |  |
| 65. | How did your current provider communicate the results of the sputum test with you? | a. In person at the health facility b. They called me c. They sent me a text message d. They sent me an e-mail e. Other, specify:__________ |  |
| 66. | How much time passed between when you were told you had TB and when you first started taking your TB medication?  **Read answer options** | a. Same day or up to 2 days after diagnosis b. 3-7 days after diagnosis (up to 1 week) c. More than 1 week but less than 1 month after diagnosis d. 1 to 2 months after diagnosis e. Over 2 months after diagnosis |  |
| 67. | How were you instructed to collect your medication?  **Read answer options** | a. Visit THIS facility to collect medication regularly b. Visit a DIFFERENT clinical facility to collect medication regularly c. Visit a pharmacy or drug shop to collect medication regularly d. Someone comes to your house to drop off the medication regularly e. Other, specify: ______ |  |
| 68. | Please select the statement that best describes your accessibility to TB treatment  **Read answer options** | a. I have no challenges accessing my TB medications b. I have some challenges accessing my TB medications c. I have many challenges accessing my TB medications | If a, skip to 65. |
| 69. | What challenges do you face when accessing your TB medications?  **Read answer options and check all that apply** | a. Not being able to get my medicines on time/when I need them b. Being exposed to COVID-19 c. People finding out I am on TB treatment d. Drug side effects e. High or unpredictable costs of treatment f. Transportation challenges g. Other, specify: ______ |  |
| **Section 6: Costing section** | | | |
| 70. | How do your current healthcare costs in general compare to your costs before the COVID pandemic started in March 2020?  **Read answer options and check one** | a. Costs decreased b. Costs stayed the same c. Costs increased d. Prefer not to say e. I do not know |  |
| 71. | What is the consultation fee at **this** health facility? | a. Price: ______ (put ‘0’ if you were not charged) b. I do not know c. Prefer not to say |  |
| 72. | What is the price you have to pay for lab tests at this facility? | a. Price: ______ (put ‘0’ if you were not charged) b. I do not know c. Prefer not to say d. N/A |  |
| 73. | What is the price of TB medication currently, per month? | a. Price: ______ (put ‘0’ if you were not charged) b. I am not on TB medications c. I do not know d. Prefer not to say |  |

**Table B: Onset of symptoms and care-seeking pathway.**

| **Onset of symptoms and care-seeking pathway** | **Patna** | **Mumbai** |
| --- | --- | --- |
|  | **Total (N =200)** | **Total (N =200)** |
| **Type of illness/symptom that led the patient to seek care at this facility** |  |  |
| Cough | 175 (87.5%) | 121 (60.5%) |
| Chest congestion | 93 (46.5%) | 39 (19.5%) |
| Head congestion | 50 (25.0%) | 20 (10.0%) |
| Sore throat | 72 (36.0%) | 18 (9.0%) |
| Rashes or allergies | 14 (7.0%) | 3 (1.5%) |
| Trouble breathing/wheezing | 90 (45.0%) | 44 (22.0%) |
| Weight loss | 154 (77.0%) | 92 (46.0%) |
| Night sweats | 59 (29.5%) | 15 (7.5%) |
| Fever | 155 (77.5%) | 124 (62.0%) |
| Lack of appetite | 143 (71.5%) | 85 (42.5%) |
| Other | 68 (34.0%) | 128 (64.0%) |
| **Before coming to this facility, whether patient sought care from any other healthcare** |  |  |
| **No** | 58 (29.0%) | 17 (8.5%) |
| **Yes** | 142 (71.0%) | 179 (89.5%) |
| **Missing** |  | 4 (2.0%) |
| **Before coming to this facility how many any other healthcare patient sought care** |  |  |
| Mean (SD) | 1.15 (1.12) | 1.69 (0.92) |
| Median [Min, Max] | 1.00 [0, 7.00] | 1.00 [0, 5.00] |
| Missing |  | 18 (9.0%) |
| **Which provider did the patient go to first to seek care?** |  |  |
| Community health worker/CHEW | 0 (0%) | 1 (0.5%) |
| Community pharmacy, chemist, drug shop, patent medicine vendor (PPMV) | 56 (28.0%) | 2 (1.0%) |
| Public outpatient clinic (e.g., PHC /Health Post/Dispensary) | 1 (0.5%) | 2 (1.0%) |
| Public Hospital | 19 (9.5%) | 4 (2.0%) |
| Private outpatient clinic, health centre, or dispensary | 62 (31.0%) | 150 (75.0%) |
| Private hospital or nursing home | 3 (1.5%) | 20 (10.0%) |
| Emergency room in a public facility | 0 (0%) | 0 (0%) |
| Emergency room in a private facility | 0 (0%) | 0 (0%) |
| Traditional healers | 0 (0%) | 0 (0%) |
| Other | 1 (0.5%) | 0 (0%) |
| Missing | 58 (29.0%) | 21 (10.5%) |
| **Which provider did the patient go to second to seek care?** |  |  |
| Community health worker/CHEW | 0 (0%) | 0 (0%) |
| Community pharmacy, chemist, drug shop, patent medicine vendor (PPMV) | 1 (0.5%) | 3 (1.5%) |
| Public outpatient clinic (e.g., PHC /Health Post/Dispensary) | 0 (0%) | 0 (0%) |
| Public Hospital | 7 (3.5%) | 9 (4.5%) |
| Private outpatient clinic, health centre, or dispensary | 41 (20.5%) | 51 (25.5%) |
| Private hospital or nursing home | 2 (1.0%) | 21 (10.5%) |
| Emergency room in a public facility | 0 (0%) | 0 (0%) |
| Emergency room in a private facility | 0 (0%) | 0 (0%) |
| Traditional healers | 0 (0%) | 0 (0%) |
| Other | 2 (1.0%) | 0 (0%) |
| Missing | 147 (73.5%) | 116 (58.0%) |
| **Which provider did the patient go to third to seek care?** |  |  |
| Community health worker/CHEW | 0 (0%) | 0 (0%) |
| Community pharmacy, chemist, drug shop, patent medicine vendor (PPMV) | 1 (0.5%) | 0 (0%) |
| Public outpatient clinic (e.g., PHC /Health Post/Dispensary) | 0 (0%) | 0 (0%) |
| Public Hospital | 5 (2.5%) | 6 (3.0%) |
| Private outpatient clinic, health centre, or dispensary | 15 (7.5%) | 12 (6.0%) |
| Private hospital or nursing home | 2 (1.0%) | 14 (7.0%) |
| Emergency room in a public facility | 0 (0%) | 0 (0%) |
| Emergency room in a private facility | 0 (0%) | 0 (0%) |
| Traditional healers | 0 (0%) | 0 (0%) |
| Other | 0 (0%) | 0 (0%) |
| Missing | 177 (88.5%) | 168 (84.0%) |
| **Which provider did the patient go to fourth to seek care?** |  |  |
| Community health worker/CHEW | 0 (0%) | 0 (0%) |
| Community pharmacy, chemist, drug shop, patent medicine vendor (PPMV) | 0 (0%) | 0 (0%) |
| Public outpatient clinic (e.g., PHC /Health Post/Dispensary) | 0 (0%) | 0 (0%) |
| Public Hospital | 1 (0.5%) | 1 (0.5%) |
| Private outpatient clinic, health centre, or dispensary | 5 (2.5%) | 7 (3.5%) |
| Private hospital or nursing home | 2 (1.0%) | 0 (0%) |
| Emergency room in a public facility | 0 (0%) | 0 (0%) |
| Emergency room in a private facility | 0 (0%) | 0 (0%) |
| Traditional healers | 0 (0%) | 0 (0%) |
| Other | 0 (0%) | 0 (0%) |
| Missing | 192 (96.0%) | 192 (96.0%) |
| **Which provider did the patient go to fifth to seek care?** |  |  |
| Community health worker/CHEW | 0 (0%) | 0 (0%) |
| Community pharmacy, chemist, drug shop, patent medicine vendor (PPMV) | 0 (0%) | 0 (0%) |
| Public outpatient clinic (e.g., PHC /Health Post/Dispensary) | 0 (0%) | 0 (0%) |
| Public Hospital | 1 (0.5%) | 0 (0%) |
| Private outpatient clinic, health centre, or dispensary | 1 (0.5%) | 1 (0.5%) |
| Private hospital or nursing home | 0 (0%) | 1 (0.5%) |
| Emergency room in a public facility | 0 (0%) | 0 (0%) |
| Emergency room in a private facility | 0 (0%) | 0 (0%) |
| Traditional healers | 0 (0%) | 0 (0%) |
| Other | 0 (0%) | 0 (0%) |
| Missing | 198 (99.0%) | 198 (99.0%) |
| **Lab test recommended by various providers** |  |  |
| First healthcare provider | 64 (32.0%) | 120 (60.0%) |
| Second healthcare provider | 39 (19.5%) | 63 (31.5%) |
| Third healthcare provider | 17 (8.5%) | 24 (12.0%) |
| Fourth healthcare provider | 7 (3.5%) | 4 (2.0%) |
| Fifth healthcare provider | 1 (0.5%) | 2 (1.0%) |
| **Types of lab test recommended by first provider** |  |  |
| Sputum test (e.g., testing mucus/phlegm using smear microscopy, AFB, GenXpert, MCS) | 32 (16.0%) | 18 (9.0%) |
| Chest X-ray | 15 (7.5%) | 22 (11.0%) |
| COVID-test (nasal/oral swab | 10 (5.0%) | 7 (3.5%) |
| Blood test | 3 (1.5%) | 0 (0%) |
| Unknown |  | 1 (0.5%) |
| Other |  |  |
| **Types of lab test recommended by second provider** |  |  |
| Sputum test (e.g., testing mucus/phlegm using smear microscopy, AFB, GenXpert, MCS) | 47 (23.5%) | 78 (39.0%) |
| Chest X-ray | 30 (15.0%) | 40 (20.0%) |
| COVID-test (nasal/oral swab | 13 (6.5%) | 16 (8.0%) |
| Blood test | 6 (3.0%) | 4 (2.0%) |
| Unknown |  | 1 (0.5%) |
| Other |  |  |
| **Types of lab test recommended by third provider** |  |  |
| Sputum test (e.g., testing mucus/phlegm using smear microscopy, AFB, GenXpert, MCS) | 15 (7.5%) | 6 (3.0%) |
| Chest X-ray | 6 (3.0%) | 4 (2.0%) |
| COVID-test (nasal/oral swab | 5 (2.5%) | 5 (2.5%) |
| Blood test | 1 (0.5%) | 0 (0%) |
| Unknown |  | 0 (0 %) |
| Other |  |  |
| **Types of lab test recommended by fourth provider** |  |  |
| Sputum test (e.g., testing mucus/phlegm using smear microscopy, AFB, GenXpert, MCS) | 55 (27.5%) | 82 (41.0%) |
| Chest X-ray | 30 (15.0%) | 45 (22.5%) |
| COVID-test (nasal/oral swab | 16 (8.0%) | 21 (10.5%) |
| Blood test | 4 (2.0%) | 3 (1.5%) |
| Unknown |  | 1 (0.5%) |
| Other |  |  |
| **Types of lab test recommended by fifth provider** |  |  |
| Sputum test (e.g., testing mucus/phlegm using smear microscopy, AFB, GenXpert, MCS) | 0 (0%) | 0 (0%) |
| Chest X-ray | 0 (0%) | 0 (0%) |
| COVID-test (nasal/oral swab | 0 (0%) | 0 (0%) |
| Blood test | 0 (0%) | 0 (0%) |
| Unknown |  |  |
| Other |  |  |
| **Patients were able to get the tests done as recommended the by first healthcare provider** |  |  |
| Yes, with no difficulties | 59 (29.5%) | 111 (55.5%) |
| Yes, with difficulties | 4 (2.0%) | 2 (1.0%) |
| No, lab was closed | 0 (0%) | 0 (0%) |
| No, I couldn’t produce sputum | 0 (0%) | 2 (1.0%) |
| No, I didn’t have time | 0 (0%) | 0 (0%) |
| Don’t know/don’t remember | 0 (0%) | 0 (0%) |
| Other | 1 (0.5%) | 0 (0%) |
| Missing | 136 (68.0%) | 85 (42.5%) |
| **Patients were able to get the tests done recommended by second healthcare provider** |  |  |
| Yes, with no difficulties | 36 (18.0%) | 55 (27.5%) |
| Yes, with difficulties | 1 (0.5%) | 1 (0.5%) |
| No, lab was closed | 0 (0%) | 0 (0%) |
| No, I couldn’t produce sputum | 1 (0.5%) | 1 (0.5%) |
| No, I didn’t have time | 0 (0%) | 1 (0.5%) |
| Don’t know/don’t remember | 0 (0%) | 0 (0%) |
| Other | 1 (0.5%) | 1 (0.5%) |
| Missing | 161 (80.5%) | 141 (70.5%) |
| **Patients were able to get the tests done recommended by third healthcare provider** |  |  |
| Yes, with no difficulties | 16 (8.0%) | 21 (10.5%) |
| Yes, with difficulties | 1 (0.5%) | 0 (0%) |
| No, lab was closed | 0 (0%) | 0 (0%) |
| No, I couldn’t produce sputum | 0 (0%) | 1 (0.5%) |
| No, I didn’t have time | 0 (0%) | 0 (0%) |
| Don’t know/don’t remember | 0 (0%) | 0 (0%) |
| Other | 0 (0%) | 0 (0%) |
| Missing | 183 (91.5%) | 178 (89.0%) |
| **Patients were able to get the tests done recommended by fourth healthcare provider** |  |  |
| Yes, with no difficulties | 7 (3.5%) | 3 (1.5%) |
| Yes, with difficulties | 0 (0%) | 0 (0%) |
| No, lab was closed | 0 (0%) | 0 (0%) |
| No, I couldn’t produce sputum | 0 (0%) | 0 (0%) |
| No, I didn’t have time | 0 (0%) | 1 (0.5%) |
| Don’t know/don’t remember | 0 (0%) | 0 (0%) |
| Other | 0 (0%) | 0 (0%) |
| Missing | 193 (96.5%) | 196 (98.0%) |
| **Sputum collected place when Sputum test was recommended by first healthcare provider** |  |  |
| At a private clinic/testing centre/ laboratory | 15 (7.5%) | 8 (4.0%) |
| At a public facility/testing centre/laboratory | 15 (7.5%) | 9 (4.5%) |
| At a mobile testing centre/camps Sputum collected at home | 1 (0.5%) | 0 (0%) |
| Sputum test recommended, but patient did not follow through with the test | 0 (0%) | 0 (0%) |
| Other | 1 (0.5%) | 0 (0%) |
| Missing | 168 (84.0%) | 183 (91.5%) |
| **Sputum collected place when Sputum test was recommended by second healthcare provider** |  |  |
| At a private clinic/testing centre/ laboratory | 1 (0.5%) | 10 (5.0%) |
| At a public facility/testing centre/laboratory | 13 (6.5%) | 11 (5.5%) |
| At a mobile testing centre/camps Sputum collected at home | 0 (0%) | 0 (0%) |
| Sputum test recommended, but patient did not follow through with the test | 0 (0%) | 0 (0%) |
| Other | 0 (0%) | 0 (0%) |
| Missing | 186 (93.0%) | 179 (89.5%) |
| **Sputum collected place when Sputum test was recommended by third healthcare provider** |  |  |
| At a private clinic/testing centre/ laboratory | 4 (2.0%) | 3 (1.5%) |
| At a public facility/testing centre/laboratory | 6 (3.0%) | 4 (2.0%) |
| At a mobile testing centre/camps Sputum collected at home | 0 (0%) | 0 (0%) |
| Sputum test recommended, but patient did not follow through with the test | 0 (0%) | 0 (0%) |
| Other | 0 (0%) | 0 (0%) |
| Missing | 190 (95.0%) | 193 (96.5%) |
| **Sputum collected place when Sputum test was recommended by fourth healthcare provider** |  |  |
| At a private clinic/testing centre/ laboratory | 0 (0%) | 0 (0%) |
| At a public facility/testing centre/laboratory | 3 (1.5%) | 1 (0.5%) |
| At a mobile testing centre/camps Sputum collected at home | 0 (0%) | 0 (0%) |
| Sputum test recommended, but patient did not follow through with the test | 0 (0%) | 0 (0%) |
| Other | 0 (0%) | 0 (0%) |
| Missing | 197 (98.5%) | 199 (99.5%) |
| **Duration of receiving the sputum test result from first provider** |  |  |
| Same day, or 1-2 days | 13 (6.5%) | 12 (6.0%) |
| 3-7 days (up to 1 week) | 14 (7.0%) | 4 (2.0%) |
| More than 1 week but less than 1 month | 3 (1.5%) | 0 (0%) |
| 1-2 months | 0 (0%) | 0 (0%) |
| 2-3 months | 0 (0%) | 0 (0%) |
| Over 3 months | 0 (0%) | 0 (0%) |
| Never received my result | 1 (0.5%) | 0 (0%) |
| Missing | 169 (84.5%) | 184 (92.0%) |
| **Duration of receiving the sputum test result from second provider** |  |  |
| Same day, or 1-2 days | 3 (1.5%) | 7 (3.5%) |
| 3-7 days (up to 1 week) | 9 (4.5%) | 12 (6.0%) |
| More than 1 week but less than 1 month | 1 (0.5%) | 1 (0.5%) |
| 1-2 months | 0 (0%) | 0 (0%) |
| 2-3 months | 0 (0%) | 0 (0%) |
| Over 3 months | 0 (0%) | 0 (0%) |
| Never received my result | 1 (0.5%) | 0 (0%) |
| Missing | 186 (93.0%) | 180 (90.0%) |
| **Duration of receiving the sputum test result from third provider** |  |  |
| Same day, or 1-2 days | 2 (1.0%) | 7 (3.5%) |
| 3-7 days (up to 1 week) | 6 (3.0%) | 0 (0%) |
| More than 1 week but less than 1 month | 1 (0.5%) | 0 (0%) |
| 1-2 months | 0 (0%) | 0 (0%) |
| 2-3 months | 0 (0%) | 0 (0%) |
| Over 3 months | 0 (0%) | 0 (0%) |
| Never received my result | 1 (0.5%) | 0 (0%) |
| Missing | 190 (95.0%) | 193 (96.5%) |
| **Duration of receiving the sputum test result from fourth provider** |  |  |
| Same day, or 1-2 days | 2 (1.0%) | 0 (0%) |
| 3-7 days (up to 1 week) | 1 (0.5%) | 0 (0%) |
| More than 1 week but less than 1 month | 0 (0%) | 0 (0%) |
| 1-2 months | 0 (0%) | 0 (0%) |
| 2-3 months | 0 (0%) | 0 (0%) |
| Over 3 months | 0 (0%) | 0 (0%) |
| Never received my result | 0 (0%) | 0 (0%) |
| Missing | 197 (98.5%) | 200 (100%) |
| **How did the first provider communicate patient with the sputum test result** |  |  |
| In person at the health facility | 27 (13.5%) | 16 (8.0%) |
| They called me | 0 (0%) | 0 (0%) |
| They sent me a text message | 0 (0%) | 0 (0%) |
| They sent me an e-mail | 0 (0%) | 0 (0%) |
| Other | 3 (1.5%) | 0 (0%) |
| Missing | 170 (85.0%) | 184 (92.0%) |
| **How did the second provider communicate patient with the sputum test result** |  |  |
| In person at the health facility | 13 (6.5%) | 16 (8.0%) |
| They called me | 0 (0%) | 0 (0%) |
| They sent me a text message | 0 (0%) | 2 (1.0%) |
| They sent me an e-mail | 0 (0%) | 0 (0%) |
| Other | 0 (0%) | 1 (0.5%) |
| Missing | 187 (93.5%) | 181 (90.5%) |
| **How did the third provider communicate patient with the sputum test result** |  |  |
| In person at the health facility | 8 (4.0%) | 7 (3.5%) |
| They called me | 1 (0.5%) | 0 (0%) |
| They sent me a text message | 0 (0%) | 0 (0%) |
| They sent me an e-mail | 0 (0%) | 0 (0%) |
| Other | 1 (0.5%) | 0 (0%) |
| Missing | 190 (95.0%) | 193 (96.5%) |
| **How did the fourth provider communicate patient with the sputum test result** |  |  |
| In person at the health facility | 3 (1.5%) | 0 (0%) |
| They called me | 0 (0%) | 0 (0%) |
| They sent me a text message | 0 (0%) | 0 (0%) |
| They sent me an e-mail | 0 (0%) | 0 (0%) |
| Other | 0 (0%) | 0 (0%) |
| Missing | 197 (98.5%) | 200 (100%) |
| **Duration to go next provider after visited first provider** |  |  |
| 1 or 2 days | 11 (5.5%) | 21 (10.5%) |
| 3-7 days (up to 1 week) | 27 (13.5%) | 57 (28.5%) |
| More than 1 week but less than 1 month | 56 (28.0%) | 73 (36.5%) |
| 1-2 months | 27 (13.5%) | 14 (7.0%) |
| 2-3 months | 3 (1.5%) | 7 (3.5%) |
| Over 3 months | 18 (9.0%) | 7 (3.5%) |
| Missing | 58 (29.0%) | 21 (10.5%) |
| **Duration to go next provider after visited second provider** |  |  |
| 1 or 2 days | 2 (1.0%) | 13 (6.5%) |
| 3-7 days (up to 1 week) | 13 (6.5%) | 37 (18.5%) |
| More than 1 week but less than 1 month | 11 (5.5%) | 27 (13.5%) |
| 1-2 months | 18 (9.0%) | 2 (1.0%) |
| 2-3 months | 2 (1.0%) | 3 (1.5%) |
| Over 3 months | 7 (3.5%) | 3 (1.5%) |
| Missing | 147 (73.5%) | 115 (57.5%) |
| **Duration to go next provider after visited third provider** |  |  |
| 1 or 2 days | 1 (0.5%) | 4 (2.0%) |
| 3-7 days (up to 1 week) | 6 (3.0%) | 13 (6.5%) |
| More than 1 week but less than 1 month | 5 (2.5%) | 8 (4.0%) |
| 1-2 months | 6 (3.0%) | 4 (2.0%) |
| 2-3 months | 1 (0.5%) | 1 (0.5%) |
| Over 3 months | 5 (2.5%) | 1 (0.5%) |
| Missing | 176 (88.0%) | 169 (84.5%) |
| **Duration to go next provider after visited fourth provider** |  |  |
| 1 or 2 days | 0 (0%) | 1 (0.5%) |
| 3-7 days (up to 1 week) | 1 (0.5%) | 1 (0.5%) |
| More than 1 week but less than 1 month | 3 (1.5%) | 3 (1.5%) |
| 1-2 months | 1 (0.5%) | 1 (0.5%) |
| 2-3 months | 0 (0%) | 1 (0.5%) |
| Over 3 months | 3 (1.5%) | 1 (0.5%) |
| Missing | 192 (96.0%) | 192 (96.0%) |
| **Place where the patients were diagnosed with TB** |  |  |
| Community health worker/CHEW | 0 (0%) | 0 (0%) |
| Community pharmacy, chemist, drug shop, patent medicine vendor (PPMV) | 1 (0.5%) | 1 (0.5%) |
| Public outpatient clinic (e.g., PHC /Health Post/Dispensary) | 0 (0%) | 0 (0%) |
| Public Hospital | 22 (11.0%) | 2 (1.0%) |
| Private outpatient clinic, health centre, or dispensary | 172 (86.0%) | 86 (43.0%) |
| Private hospital or nursing home | 5 (2.5%) | 107 (53.5%) |
| Emergency room in a public facility | 0 (0%) | 0 (0%) |
| Emergency room in a private facility | 0 (0%) | 0 (0%) |
| Traditional healers | 0 (0%) | 0 (0%) |
| Other | 0 (0%) | 0 (0%) |
| Missing |  | 4 (2.0%) |
| **TB diagnosis place and first health facility where the patient went after noticing symptom are same places** |  |  |
| No | 105 (52.5%) | 144 (72.0%) |
| Yes | 95 (47.5%) | 52 (26.0%) |
| Missing |  | 4 (2.0%) |
| **Place where the patients received TB treatment** |  |  |
| Community health worker/CHEW | 1 (0.5%) | 2 (1.0%) |
| Community pharmacy, chemist, drug shop, patent medicine vendor (PPMV) | 0 (0%) | 0 (0%) |
| Public outpatient clinic (e.g., PHC /Health Post/Dispensary) | 0 (0%) | 0 (0%) |
| Public Hospital | 24 (12.0%) | 5 (2.5%) |
| Private outpatient clinic, health centre, or dispensary | 170 (85.0%) | 60 (30.0%) |
| Private hospital or nursing home | 5 (2.5%) | 129 (64.5%) |
| Emergency room in a public facility | 0 (0%) | 0 (0%) |
| Emergency room in a private facility | 0 (0%) | 0 (0%) |
| Traditional healers | 0 (0%) | 0 (0%) |
| Other | 0 (0%) | 0 (0%) |
| Missing |  | 4 (2.0%) |
| **TB treatment place and the health facility where the patient was diagnosed are same places** |  |  |
| No | 53 (26.5%) | 76 (38.0%) |
| Yes | 147 (73.5%) | 120 (60.0%) |
| Missing |  | 4 (2.0%) |
| **The current provider recommended lab test** |  |  |
| No | 6 (3.0%) | 5 (2.5%) |
| Yes | 194 (97.0%) | 190 (95.0%) |
| Missing |  | 5 (2.5%) |
| **Types of lab test recommended by the current provider** |  |  |
| Sputum test (e.g. smear microscopy, AFB, GenXpert, MCS) | 158 (79.0%) | 131 (65.5%) |
| Chest X-ray | 184 (92.0%) | 136 (68.0%) |
| COVID-test | 34 (17.0%) | 19 (9.5%) |
| HIV test | 54 (27.0%) | 55 (27.5%) |
| Malaria test | 5 (2.5%) | 8 (4.0%) |
| Typhoid test | 11 (5.5%) | 18 (9.0%) |
| Blood test | 189 (94.5%) | 158 (79.0%) |
| Unknown | 2 (1.0%) | 2 (1.0%) |
| Other | 63 (31.5%) | 124 (62.0%) |
| **Recommended tests by the current provider were done** |  |  |
| Yes, with no difficulties | 184 (92.0%) | 188 (94.0%) |
| Yes, with difficulties | 5 (2.5%) | 0 (0%) |
| No, lab was closed | 0 (0%) | 0 (0%) |
| No, I couldn't produce sputum | 4 (2.0%) | 0 (0%) |
| No, I didn't have time | 1 (0.5%) | 1 (0.5%) |
| Don't know/don't remember | 0 (0%) | 0 (0%) |
| Other | 0 (0%) | 0 (0%) |
| Missing | 6 (3.0%) | 11 (5.5%) |
| **Sputum test result collected place recommended by current provider** | |  |
| Here in this facility | 61 (30.5%) | 72 (36.0%) |
| At a public facility/testing centre/laboratory | 81 (40.5%) | 59 (29.5%) |
| At a mobile testing centre/camps | 4 (2.0%) | 0 (0%) |
| Sputum collected at home | 7 (3.5%) | 0 (0%) |
| Sputum test recommended, but patient did not follow through with the test | 0 (0%) | 0 (0%) |
| Other | 1 (0.5%) | 0 (0%) |
| Missing | 46 (23.0%) | 69 (34.5%) |
| **How did the current provider communicate patient with the sputum test result** |  |  |
| In person at the health facility | 145 (72.5%) | 128 (64.0%) |
| They called me | 1 (0.5%) | 0 (0%) |
| They sent me a text message | 0 (0%) | 1 (0.5%) |
| They sent me an e-mail | 0 (0%) | 0 (0%) |
| Other | 8 (4.0%) | 3 (1.5%) |
| Missing | 46 (23.0%) | 68 (34.0%) |
| **How patients were instructed to collect medication by current provider** | |  |
| Visit THIS facility to collect medication regularly | 64 (32.0%) | 78 (39.0%) |
| Visit a DIFFERENT clinical facility to collect medication regularly | 6 (3.0%) | 28 (14.0%) |
| Visit a pharmacy or drug shop to collect medication regularly | 128 (64.0%) | 84 (42.0%) |
| Someone comes to your house to drop off the medication regularly | 0 (0%) | 0 (0%) |
| Other | 2 (1.0%) | 6 (3.0%) |
| Missing |  | 4 (2.0%) |
| **Patient accessibility to TB treatment** |  |  |
| I have no challenges accessing my TB medications | 178 (89.0%) | 194 (97.0%) |
| I have some challenges accessing my TB medications | 15 (7.5%) | 2 (1.0%) |
| I have many challenges accessing my TB medications | 7 (3.5%) | 0 (0%) |
| Missing |  | 4 (2.0%) |
| **Challenges faced by patient when accessing TB medications** |  |  |
| Not being able to get my medicines on time/when I need them | 16 (8.0%) | 2 (1.0%) |
| Being exposed to COVID-19 | 0 (0%) | 0 (0%) |
| People finding out I am on TB treatment | 1 (0.5%) | 2 (1.0%) |
| Drug side effects | 2 (1.0%) | 0 (0%) |
| High or unpredictable costs of treatment | 5 (2.5%) | 0 (0%) |
| Transportation challenges | 6 (3.0%) | 0 (0%) |
| Other | 2 (1.0%) | 0 (0%) |
| **No of encounters** |  |  |
| Mean (SD) | 2.15 (1.12) | 2.69 (0.920) |
| Median [Min, Max] | 2.00 [1.00, 8.00] | 2.00 [1.00, 6.00] |
| Missing |  | 18 (9.0%) |

**Table C: Perspectives on the COVID-19 pandemic.**

| Perspectives on the COVID-19 pandemic | Patna | Mumbai |
| --- | --- | --- |
|  | **Total (N =200)** | **Total (N =200)** |
| Any COVID lockdown around the residence |  |  |
| No | 0 (0%) | 0 (0%) |
| Yes | 200 (100%) | 195 (97.5%) |
| Missing |  | 5 (2.5%) |
| Types of COVID related restriction |  |  |
| Movement completely restricted | 2 (1%) | 0 (0%) |
| Restaurants/bar open with limited capacity | 0 (0%) | 0 (0%) |
| Markets/stores open with limited capacity |  | 0 (0%) |
| Religious ceremonies/services/mosques at limited capacity | 1 (0.5%) | 0 (0%) |
| Movement partially restricted | 1 (0.5%) | 0 (0%) |
| Masks mandatory in public spaces | 85 (42.5%) | 0 (0%) |
| Social distance mandated | 75 (37.5%) | 0 (0%) |
| Large gathering restricted | 26 (13.0%) | 0 (0%) |
| Others restriction | 88 (44.0%) | 195 (97.5%) |
| How COVID-related lockdown/restriction affected Access to healthcare |  |  |
| Unable to reach a doctor because facilities were closed | 29 (14.5%) | 8 (4.0%) |
| Unable to receive medication because drug stores were closed | 8 (4.0%) | 1 (0.5%) |
| Obliged to seek care from a different facility than preferred | 7 (3.5%) | 3 (1.5%) |
| Waiting time became longer than normal/expected | 19 (9.5%) | 43 (21.5%) |
| Unable to leave the house to seek care because of movement restriction | 24 (12.0%) | 16 (8.0%) |
| Other affect on Health care | 4 (2.0%) | 132 (66.0%) |
| Friends/family members diagnosed with COVID |  |  |
| No | 175 (87.5%) | 160 (80.0%) |
| Yes | 25 (12.5%) | 33 (16.5%) |
| Missing |  | 7 (3.5%) |
| COVID and current/past COVID-related restrictions affect willingness to seek healthcare |  |  |
| Yes, I am more willing to seek care | 63 (31.5%) | 101 (50.5%) |
| Yes, I am less willing to seek care | 19 (9.5%) | 10 (5.0%) |
| No effect | 118 (59.0%) | 83 (41.5%) |
| Don't know/not sure | 0 (0%) | 0 (0%) |
| Missing |  | 6 (3.0%) |
| How COVID has decreased willingness to seek healthcare |  |  |
| Creates fear of infection while visiting health facilities | 17 (8.5%) | 7 (3.5%) |
| Creates fear of being diagnosed with COVID while visiting health facilities | 7 (3.5%) | 4 (2.0%) |
| Made inability to seek care due to loss of income | 6 (3.0%) | 3 (1.5%) |
| Created other factors that decreased willingness to seek care | 0 (0%) | 0 (0%) |
| How COVID has increased willingness to seek healthcare |  |  |
| Increased awareness to seek early health care | 58 (29.0%) | 100 (50.0%) |
| Increased awareness to get tested regularly for work/school | 5 (2.5%) | 1 (0.5%) |
| Increased frequent healthcare seeking due to awareness about family members’ infection | 26 (13.0%) | 6 (3.0%) |
| Increased frequent healthcare seeking due to awareness about getting infected by other people | 17 (8.5%) | 6 (3.0%) |
| Increased frequent healthcare seeking due to awareness about staying with family members | 13 (6.5%) | 2 (1.0%) |
| Increased willingness to healthcare seeking due to understanding of underlying/chronic condition | 17 (8.5%) | 2 (1.0%) |
| Increased other awareness to seek health care | 1 (0.5%) | 1 (0.5%) |

**Table D: Health Care Utilization during the COVID-19 Pandemic.**

| **Health Care Utilization during the COVID-19 pandemic** | **Patna** | **Mumbai** |
| --- | --- | --- |
|  | **Total (N =200)** | **Total (N =200)** |
| **Types of health providers who were contacted since the beginning of the COVID-19 pandemic for physical or mental health** |  |  |
| Community health worker/CHEW | 3 (1.5%) | 0 (0%) |
| Community pharmacy, chemist, drug shop, patent medicine vendor (PPMV) | 105 (52.5%) | 3 (1.5%) |
| Private laboratory | 34 (17.0%) | 1 (0.5%) |
| Public outpatient clinic (e.g., PHC /Health Post/Dispensary) | 5 (2.5%) | 5 (2.5) |
| Public Hospital | 59 (29.5%) | 44 (22.0%) |
| Private outpatient clinic, health centre, or dispensary | 181 (90.5%) | 182 (91.0%) |
| Private hospital or nursing home | 6 (3.0%) | 120 (60.0%) |
| Emergency room in a public facility | 4 (2.0%) | 1 (0.5%) |
| Emergency room in a private facility | 2 (1.0%) | 2 (1.0%) |
| Traditional healers | 3 (1.5%) | 0 (0%) |
| Other | 4 (2.0%) | 0 (0%) |
| Missing |  | 4(2.0%) |
| **Place to go to seek help first to get treatment for sickness** |  |  |
| Community health worker/CHEW | 1 (0.5%) | 0 (0%) |
| Community pharmacy, chemist, drug shop, patent medicine vendor (PPMV) | 82 (41.0%) | 3 (1.5%) |
| Private laboratory | 0 (0%) | 0 (0%) |
| Public outpatient clinic (e.g., PHC /Health Post/Dispensary) | 0 (0%) | 1 (0.5%) |
| Public Hospital | 12 (6.0%) | 2 (1.0%) |
| Private outpatient clinic, health centre, or dispensary | 103 (51.5%) | 184 (92.0%) |
| Private hospital or nursing home | 2 (1.0%) | 5 (2.5%) |
| Emergency room in a public facility | 0 (0%) | 0 (0%) |
| Emergency room in a private facility | 0 (0%) | 0 (0%) |
| Traditional healers | 0 (0%) | 0 (0%) |
| Other | 0 (0%) | 0 (0%) |
| Missing |  | 5(2.5%) |
| **Patient felt the necessity of healthcare but did not receive it** |  |  |
| No | 181 (90.5%) | 191 (95.5%) |
| Yes | 19 (9.5%) | 5 (2.5%) |
| Can't remember | 0 (0%) | 0 (0%) |
| Missing |  | 4(2.0%) |
| **Reason for not receiving healthcare even if it was needed** |  |  |
| Healthcare was not available because practice was closed due to the COVID-19 pandemic | 13 (6.5%) | 3 (1.5%) |
| Healthcare was not available because practice was closed, but not due to COVID-19 | 2 (1.0%) | 2 (1.0%) |
| Waiting time was too long | 0 (0%) | 0 (0%) |
| Could not afford | 4 (2.0%) | 0 (0%) |
| Too busy with work and other responsibilities | 0 (0%) | 0 (0%) |
| Decided not to seek care due to fear of exposure to COVID-19 pandemic | 0 (0%) | 1 (0.5%) |
| Other | 3 (1.5%) | 0 (0.5%) |
| **Before the pandemic frequency of using telemedicine to consult with healthcare professionals** |  |  |
| Did not use telemedicine | 191 (95.5%) | 194 (97.0%) |
| Daily | 0 (0%) | 0 (0%) |
| More than once a week | 1 (0.5%) | 0 (0%) |
| Once a week | 1 (0.5%) | 0 (0%) |
| Less than once a week but more than once a month | 5 (2.5%) | 0 (0%) |
| Once a month or less | 2 (1.0%) | 2 (1.0%) |
| Other | 0 (0%) | 0 (0%) |
| Missing |  | 4(2.0%) |
| **Since the start of the pandemic frequency of using telemedicine to consult with healthcare professionals** |  |  |
| Did not use telemedicine | 181 (90.5%) | 188 (94.0%) |
| Daily | 2 (1.0%) | 0 (0%) |
| More than once a week | 0 (0%) | 0 (0%) |
| Once a week | 0 (0%) | 0 (0%) |
| Less than once a week but more than once a month | 5 (2.5%) | 1 (0.5%) |
| Once a month or less | 11 (5.5%) | 5 (2.5%) |
| Other | 1 (0.5%) | 2 (1.0%) |
| Missing |  | 4 (2.0%) |

**Table E: TB Diagnosis and Treatment Initiation.**

| TB Diagnosis and Treatment Initiation | Patna | Mumbai |
| --- | --- | --- |
|  | **Total (N=200)** | **Total (N=200)** |
| Ever diagnosed with TB |  |  |
| No | 0 (0%) | 0 (0%) |
| Yes | 200 (100%) | 195 (97.5%) |
| Not sure | 0 (0%) | 0 (0%) |
| Missing |  | 5 (2.5%) |
| TB care stage now |  |  |
| My doctor told me I have TB but I have not been given any medicines yet | 0 (0%) | 0 (0%) |
| I started taking TB medication, but I stopped before finishing all my medication | 15 (7.5%) | 0 (0%) |
| I am currently taking TB medication and have been taking my medicines up to today | 76 (38.0%) | 110 (55.0%) |
| I finished taking all my TB medication | 108 (54.0%) | 85 (42.5%) |
| Not sure | 1 (0.5%) | 0 (0%) |
| Other | 0 (0%) | 0 (0%) |
| Missing |  | 0 (0%) |

**Factors associated with Various delays**

**Table F: Median regression: factors associated with Health-seeking delay for Patna.**

| Characteristics |  | Unadjusted | | Adjusted | |
| --- | --- | --- | --- | --- | --- |
|  |  | **Coefficient(95% CI)** | **p-value** | **Coefficient(95% CI)** | **p-value** |
| Age |  | 0.02381 (-0.01668, 0.06430) | 0.251 | 0.02694 (-0.03305, 0.08692) | 0.38 |
| Gender | Female |  |  |  |  |
|  | Male | -1.00000 (-2.54837, 0.54837) | 0.207 | -0.79265 (-2.25760, 0.67229) | 0.29 |
| Marital Status | Single |  |  |  |  |
|  | Married | 0 (-1.97251, 1.97251) | 1 | 1.05796 (-1.23729, 3.35321) | 0.368 |
|  | Other | 0(-0.96134, 0.96134) | 1 | -0.78612 (-3.40404, 1.83179) | 0.557 |
| Number of individuals in the family |  | 0 (-0.28338, 0.28338) | 1 | 0.12490 (-0.26714, 0.51694) | 0.533 |
| Highest education level | Primary 6 or lower |  |  |  |  |
|  | Junior secondary school | 0 (-2.07886, 2.07886) | 1 | 0.20571 (-2.22018, 3.22426) | 0.848 |
|  | Senior secondary school | 0 (-2.16280, 2.16280) | 1 | -0.73143 (-3.03811, 1.57525) | 0.535 |
|  | Bachelor's degree | 2(-1.39026, 5.39026) | 0.249 | 0.50204 (-1.88958, 2.30100) | 0.718 |
|  | Post-graduate certificate | 5(-16.27974, 26.27974) | 0.646 | 3.73714 (-21.24180, 28.71609) | 0.77 |
|  | Other | 1 (-1.23548, 3.23548) | 0.382 | 0.06041 (-1.99555, 2.11637) | 0.954 |
| Employment Status | Working |  |  |  |  |
|  | Self-employed | 0 (-2.84630, 2.84630) | 1 | -0.36735 (-2.35048, 1.61578) | 0.717 |
|  | Unemployed/underemployed | 1 (-0.91624, 2.91624) | 0.308 | -0.38449 (-2.17881, 1.40983) | 0.675 |
|  | Retired | 9 (-2.22538, 20.22538) | 0.118 | 5.72327 (-5.55561, 17.00214) | 0.321 |
| Financial Standing | Adequate |  |  |  |  |
|  | Barely adequate | 1(-0.77795, 2.77795) | 0.272 | 0.20571 (-1.33460, 1.74603) | 0.794 |
|  | Inadequate | 1(-0.85822, 2.85822) | 0.293 | 0.25061 (-1.88948, 2.39070) | 0.819 |
| Contact with active Pulmonary TB people | No |  |  |  |  |
|  | Yes | 0(-1.74640, 1.74640) | 1 | -0.72735 (-2.16033, 0.70564) | 0.321 |
| Friends or family COVID diagnosed | No |  |  |  |  |
|  | Yes | 3(-1.37560, 7.37560) | 0.181 | 1.09714 (-2.37696, 4.57125) | 0.537 |
| COVID affected on willingness to seek health care | No effect |  |  |  |  |
|  | More willing | 1(-0.74419, 2.74419) | 0.263 | 0.21714 (-1.14016, 1.57445) | 0.754 |
|  | Less willing | 0(-3.06767, 3.06767) | 1 | -0.63102 (-3.62341, 2.36137) | 0.68 |

**Table G: Median regression: factors associated with Health-seeking delay for Mumbai.**

| Characteristics |  | Unadjusted | | Adjusted | |
| --- | --- | --- | --- | --- | --- |
|  |  | **Coefficient(95% CI)** | **p-value** | **Coefficient(95% CI)** | **p-value** |
| Age |  | 0.04 (-0.03,0.12) | 0.278 | 0.03 (-0.05-0.13) | 0.437 |
| Gender | Female | - |  | - |  |
|  | Male | 0.99 (-0.65,2.64) | 0.234 | 1.93 (0.13-3.72) | 0.037 |
| Marital Status | Single | - |  | - |  |
|  | Married | -0.98 (-2.52,0.51) | 0.198 | -1.18 (-2.91-0.54) | 0.182 |
|  | Other | -0.99 (-2.71-4.72) | 0.598 | 0.82 (-3.98-5.62) | 0.738 |
| Number of individuals in the family |  | 0.01 (-0.39-0.41) | 0.989 | 0.29 (-0.11-0.69) | 0.080 |
| Highest education level | Primary 6 or lower | - |  | - |  |
|  | Junior secondary school | -2.00 (-4.36-0.36) | 0.104 | -0.91 (-3.42-1.77) | 0.533 |
|  | Senior secondary school | -2.00 (-4.62-0.62) | 0.136 | -0.44 (-3.33-2.44) | 0.762 |
|  | Bachelor's degree | -2.00 (-4.20-0.21) | 0.077 | 0.02 (-2.67-2.69) | 0.989 |
|  | Post-graduate certificate | 0.01(-2.68-2.68) | 1.000 | 1.14 (-2.32-4.60) | 0.519 |
|  | Other | -1.00 (-3.39-1.39) | 0.414 | 1.51 (-3.59-4.43) | 0.839 |
| Employment Status | Working | - |  | - |  |
|  | Self-employed | 2.00 (-15.79-19.79) | 0.825 | 1.31 (-16.67-19.29) | 0.952 |
|  | Unemployed/underemployed | 0.01 (-1.69-1.70) | 0.998 | 1.19 (-0.43-2.82) | 0.152 |
|  | Retired | 3.00 (0.32-5.67) | 0.029 | 2.78 (-13.45-15.21) | 0.904 |
| Financial Standing | Adequate | - |  | - |  |
|  | Barely adequate | 0.99 (-0.73-2.73) | 0.258 | 0.19 (-1.23-1.62) | 0.794 |
|  | Inadequate | -0.99 (-9.22-7.22) | 0.812 | -2.39 (-5.68-0.89) | 0.155 |
| Contact with active Pulmonary TB people | No | - |  | - |  |
|  | Yes | 1.00 (-1.38-3.38) | 0.411 | 0.69 (-1.87-3.26) | 0.596 |
| Friends or family COVID diagnosed | No | - |  | - |  |
|  | Yes | 2.00 (0.17-3.82) | 0.033 | 1.48 (-1.73-4.67) | 0.368 |
| COVID affected on willingness to seek health care | No effect | - |  | - |  |
|  | More willing | -1.00 (-2.53-0.53) | 0.203 | -0.98 (-2.84-0.87) | 0.301 |
|  | Less willing | 1.00 (-13.07-15.07) | 0.889 | -0.86 (-20.56-18.84) | 0.931 |

**Table H: Median regression: factors associated with Provider delay in Patna.**

| **Characteristics** |  | **Unadjusted** | | **Adjusted** | |
| --- | --- | --- | --- | --- | --- |
|  |  | **Coefficient (95% CI)** | **p-value** | **Coefficient (95% CI)** | **p-value** |
| **Age** |  | 0.083 (-0.448, 0.614) | 0.759 | 0.111 (-1.015, 1.237) | 0.847 |
| **Gender** | Female |  |  |  |  |
|  | Male | -8 (-20.952, 4.952) | 0.227 | -7.917 (-43.681, 27.848) | 0.665 |
| **Marital Status** | Single |  |  |  |  |
|  | Married | -1 (-16.202, 14.202) | 0.898 | 10.750 (-27.088, 48.588) | 0.579 |
|  | Other | 22 (-2.595, 46.595) | 0.081 | 13.833 (-49.122, 76.789) | 0.667 |
| **Number of individuals in the family** |  | 0.875 (-2.105, 3.855) | 0.566 | -1.056 (-6.605, 4.494) | 0.71 |
| **Highest education level** | Primary 6 or lower |  |  |  |  |
|  | Junior secondary school | 11 (-15.667, 37.667) | 0.42 | 26.361 (-16.223, 68.945) | 0.227 |
|  | Senior secondary school | 7 (-15.341, 29.341) | 0.54 | 0.194 (-50.745, 51.134) | 0.994 |
|  | Bachelor's degree | 8 (-15.270, 31.270) | 0.501 | 1.806 (-60.714, 64.325) | 0.955 |
|  | Post-graduate certificate | -4 (-41.207, 33.207) | 0.833 | -17.278 (-113.971, 79.416) | 0.727 |
|  | Other | -4 (-26.807, 18.807) | 0.731 | 11.111 (-26.162, 48.385) | 0.56 |
| **Employment Status** | Working |  |  |  |  |
|  | Self-employed | -9 (-35.078, 17.078) | 0.5 | 0.028 (-39.349, 39.404) | 0.999 |
|  | Unemployed/underemployed | -1 (-20.576, 18.576) | 0.92 | -15.111 (-52.256, 22.034) | 0.427 |
|  | Retired | -8 (-46.754, 30.754) | 0.686 | -3.500 (-90.431, 83.431) | 0.937 |
| **Financial Standing** | Adequate |  |  |  |  |
|  | Barely adequate | 1 (-17.892, 19.892) | 0.917 | 7.639 (-23.604, 38.881) | 0.633 |
|  | Inadequate | -6 (-39.810, 27.810) | 0.728 | -0.056 (-39.879, 39.768) | 0.998 |
| **Contact with active Pulmonary TB people** | No |  |  |  |  |
|  | Yes | 3 (-13.220, 19.220) | 0.717 | 3.139 (-24.445, 30.723) | 0.824 |
| **Friends or Family COVID diagnosed** | No |  |  |  |  |
|  | Yes | -6 (-48.382, 36.382) | 0.782 | -1.583 (-82.599, 79.432) | 0.97 |
| **COVID affected on willingness to seek health care** | No effect |  |  |  |  |
|  | More willing | -10 (-24.144, 4.144) | 0.167 | 3.250 (-34.741, 41.241) | 0.867 |
|  | Less willing | 39 (-14.110, 92.110) | 0.152 | 44.028 (-33.090, 121.145) | 0.265 |
| **First health care provider** | Private outpatient clinic |  |  |  |  |
|  | Public outpatient clinic | 11 (-6.297, 28.297) | 0.215 | -2.444 (-63.766, 58.877) | 0.938 |
|  | Public Hospital | -24 (-48.882, 0.882) | 0.061 | -24.583 (-64.874, 15.708) | 0.234 |
|  | Private Hospital or nursing home | -16 (-51.147, 19.147) | 0.374 | -5.972 (-62.135, 50.190) | 0.835 |

**Table I: Median regression: factors associated with Provider delay in Mumbai.**

|  |  | **Unadjusted** | | **Adjusted** | |
| --- | --- | --- | --- | --- | --- |
| **Outcome: Provider delay** |  | **Coefficient (95% CI)** | **p-value** | **Coefficient (95% CI)** | **p-value** |
| **Age** |  | 0.16 (-0.18-0.52) | 0.358 | 0.28 (-0.39-0.96) | 0.408 |
| **Gender** | Female | - |  | - |  |
|  | Male | -0.99 (-11.38-9.38) | 0.834 | -0.55 (-10.78-9.67) | 0.915 |
| **Marital Status** | Single | - |  | - |  |
|  | Married | -4 (-13.76-5.76) | 0.423 | -3.31 (-13.09-6.47) | 0.508 |
|  | Other | -12 (-69.88-45.88) | 0.668 | -7.93 (-83.40-67.53) |  |
| **Number of individuals in the family** |  | -0.25 (-2.08-1.58) | 0.789 | -0.46 (-2.14-1.22) | 0.594 |
| **Highest education level** | Primary 6 or lower | - |  | - |  |
|  | Junior secondary school | 0.01 (-12.27-12.27) | 0.989 | 5.36 (-11.56-22.29) | 0.535 |
|  | Senior secondary school | 3 (-7.85-13.85) | 0.588 | 12.09 (-4.67-28.86) | 0.159 |
|  | Bachelor's degree | -3 (-13.44-7.44) | 0.574 | 4.52 (-13.71-22.74) | 0.511 |
|  | Post-graduate certificate | 20 (-43.34-83.34) | 0.536 | 25.21 (-36.75-87.14) | 0.426 |
|  | Other | 26 (-14.73-66.74) | 0.212 | 9.82 (-37.16-56.81) | 0.682 |
| **Employment Status** | Working | - |  | - |  |
|  | Self-employed | 3 (-27.86-33.86) | 0.849 | -2.52 (-74.25-59.62) | 0.987 |
|  | Unemployed/underemployed | 1 (-8.51-10.52) | 0.837 | 1.42 (-8.51-11.32) | 0.782 |
|  | Retired | 16 (-1.42-33.42) | 0.073 | -9.23 (-63.49-45.01) | 0.739 |
| **Financial Standing** | Adequate | - |  | - |  |
|  | Barely adequate | -1 (-9.05-7.05) | 0.808 | 6.35 (-4.14-16.84) | 0.237 |
|  | Inadequate | 2 (-31.84-35.84) | 0.970 | 1.10 (-25.37-27.58) | 0.935 |
| **Contact with active Pulmonary TB people** | No | - |  |  |  |
|  | Yes | -1 (-18.53-16.53) | 0.911 | -6.34 (-30.84-18.14) | 0.612 |
| **Friends or Family COVID diagnosed** | No |  |  |  |  |
|  | Yes | -1 (-13.73-11.74) | 0.877 | -3.88 (-18.09-10.33) | 0.592 |
| **COVID affected on willingness to seek health care** | No effect | - |  | - |  |
|  | More willing | -4 (-12.44-4.44) | 0.354 | -7.04 (-17.42-3.32) | 0.184 |
|  | Less willing | 19 (-24.29-62.29) | 0.390 | 18.42 (-39.42-76.24) | 0.533 |
| **First health care provider** | Private outpatient clinic | - |  | - |  |
|  | Public outpatient clinic | -14 (-26.16- -1.85) | 0.025 | -12 (-29.72- 4.36) | 0.871 |
|  | Public Hospital | -6 (-17.28- 5.93) | 0.325 | -10 (-17.28- 5.93) | 0.922 |
|  | Private Hospital or nursing home | -11 (-25.06-3.06) | 0.127 | -9 (-25.14-6.61) | 0.702 |

**Table J: Median regression: factors associated with Treatment delay in Patna.**

| **Characteristics** |  | **Unadjusted** | | **Adjusted** | |
| --- | --- | --- | --- | --- | --- |
|  |  | **Coefficient (95% CI)** | **p-value** | **Coefficient (95% CI)** | **p-value** |
| **Age** |  | 0.028 (-0.015, 0.070) | 0.201 | 0.020 (-0.105, 0.145) | 0.758 |
| **Gender** | Female |  |  |  |  |
|  | Male | 1 (-0.611, 2.611) | 0.225 | 2.229 (-1.683, 6.140) | 0.266 |
| **Marital Status** | Single |  |  |  |  |
|  | Married | -1 (-2.491, 0.491) | 0.19 | 0.120 (-2.731, 2.972) | 0.934 |
|  | Other | 1 (-1.382, 3.382) | 0.412 | -0.897 (-6.622, 4.829) | 0.759 |
| **Number of individuals in the family** |  | -0.222 (-0.419, -0.025) | 0.028 | -0.278 (-0.828, 0.272) | 0.324 |
| **Highest education level** | Primary 6 or lower |  |  |  |  |
|  | Junior secondary school | 1 (-1.030, 3.030) | 0.336 | 0.440 (-2.571, 3.451) | 0.775 |
|  | Senior secondary school | 0 (-8.012, 8.012) | 1 | -0.160 (-8.388, 8.068) | 0.97 |
|  | Bachelor's degree | 0 (-1.899, 1.899) | 1 | -0.120 (-3.642, 3.402) | 0.947 |
|  | Post-graduate certificate | 1 (-0.991, 2.991) | 0.326 | -0.138 (-6.119, 5.843) | 0.964 |
|  | Other | 1 (-1.252, 3.252) | 0.385 | 1.594 (-4.405, 7.594) | 0.604 |
| **Employment Status** | Working |  |  |  |  |
|  | Self-employed | 0 (-2.763, 2.763) | 1 | 1.714 (-3.336, 6.765) | 0.507 |
|  | Unemployed/underemployed | 1 (-1.074, 3.074) | 0.346 | 2.686 (-0.652, 6.024) | 0.118 |
|  | Retired | 1 (-5.585, 7.585) | 0.766 | 4.500 (-8.049, 17.050) | 0.484 |
| **Financial Standing** | Adequate |  |  |  |  |
|  | Barely adequate | 2 (0.764, 3.236) | 0.002 | 0.816 (-1.637, 3.269) | 0.516 |
|  | Inadequate | 2 (-0.359, 4.359) | 0.098 | 1.118 (-3.172, 5.408) | 0.611 |
| **Contact with active Pulmonary TB people** | No |  |  |  |  |
|  | Yes | -1 (-2.175, 0.175) | 0.097 | -0.576 (-2.953, 1.801) | 0.636 |
| **Friends or Family COVID diagnosed** | No |  |  |  |  |
|  | Yes | -1 (-2.427, 0.427) | 0.171 | -1.149 (-5.992, 3.694) | 0.643 |
| **COVID affected on willingness to seek health care** | No effect |  |  |  |  |
|  | More willing | 1 (-0.454, 2.454) | 0.179 | 0.238 (-2.759, 3.235) | 0.877 |
|  | Less willing | 1 (-10.166, 12.166) | 0.861 | 1.388 (-17.801, 20.576) | 0.888 |
| **First health care provider** | Private outpatient clinic |  |  |  |  |
|  | Public outpatient clinic | 3 (-0.006, 6.006) | 0.052 | 4.039 (-2.158, 10.235) | 0.204 |
|  | Public Hospital | 2 (-2.029, 6.029) | 0.332 | 1.033 (-4.126, 6.192) | 0.695 |
|  | Private Hospital or nursing home | 1 (-47.138, 49.138) | 0.968 | -0.725 (-41.806, 40.355) | 0.972 |
| **TB diagnosis place** | Private outpatient clinic |  |  |  |  |
|  | Public Hospital | 3 (-3.204, 9.204) | 0.344 | 2.659 (-5.146, 10.463) | 0.506 |
|  | Private hospital or nursing home | 2 (-10.656, 14.656) | 0.757 | 3.022 (-17.365, 23.409) | 0.772 |

**Table K: Median regression: factors associated with Treatment delay in Mumbai.**

|  |  | **Unadjusted** | | **Adjusted** | |
| --- | --- | --- | --- | --- | --- |
| **Outcome: Treatment delay** |  | **Coefficient (95% CI)** | **p-value** | **Coefficient (95% CI)** | **p-value** |
| **Age** |  | -0.02 (-0.04- -0.01) | 0.039 | -0.02 (-0.06-0.02) | 0.290 |
| **Gender** | Female | - |  | - |  |
|  | Male | 0.01 (-1.24-1.23) | 0.989 | 0.24 (-0.82-1.31) | 0.232 |
| **Marital Status** | Single | - |  | - |  |
|  | Married | -1.01 (-2.15-0.15) | 0.078 | -0.15 (-1.23-0.93) | 0.744 |
|  | Other | -1.01(-3.06-1.06) | 0.342 | -0.11 (-1.49-1.27) | 0.858 |
| **Number of individuals in the family** |  | -0.13 (-0.31 -0.06) | 0.204 | 0.07(-0.11-0.26) | 0.253 |
| **Highest education level** | Primary 6 or lower | - |  | - |  |
|  | Junior secondary school | 1.01 (-0.57-2.58) | 0.215 | -0.43 (-1.81-0.94) | 0.531 |
|  | Senior secondary school | 1.02 (-0.45-2.45) | 0.178 | -0.48 (-1.70-0.73) | 0.460 |
|  | Bachelor's degree | 1.01 (-0.46-2.46) | 0.180 | -0.05 (-1.20-1.09) | 0.926 |
|  | Post-graduate certificate | 0.01 (-2.12-2.12) | 0.997 | -0.67 (-2.17-0.83) | 0.362 |
|  | Other | 1.01 (-0.73-2.72) | 0.257 | 0.29 (-1.86-2.46) | 0.779 |
| **Employment Status** | Working | - |  | - |  |
|  | Self-employed | 1.02 (-23.43-25.43) | 0.936 | 1.68 (-18.81-22.19) | 0.896 |
|  | Unemployed/underemployed | 1.01 (-0.09-2.09) | 0.076 | 0.52 (-0.25-1.31) | 0.194 |
|  | Retired | 0.01 (-0.78-0.78) | 0.986 | -3.70 (-13.36-5.96) | 0.658 |
| **Financial Standing** | Adequate | - |  | - |  |
|  | Barely adequate | 1.01 (-0.49-2.49) | 0.190 | 0.28 (-0.70-1.26) | 0.561 |
|  | Inadequate | 1.01 (-0.36-2.36) | 0.150 | 0.64 (-1.72-3.01) | 0.540 |
| **Contact with active Pulmonary TB people** | No | - |  |  |  |
|  | Yes | 0.01 (-1.58-1.58) | 0.985 | -0.04 (-1.05-0.96) | 0.923 |
| **Friends or Family COVID diagnosed** | No |  |  |  |  |
|  | Yes | 1.02 (-0.64-2.64) | 0.236 | -0.33 (-1.69-1.03) | 0.636 |
| **COVID affected on willingness to seek health care** | No effect | - |  | - |  |
|  | More willing | 0.02 (-1.16-1.16) | 0.928 | 0.03 (-0.70-0.76) | 0.919 |
|  | Less willing | 1.02 (-6.33-8.33) | 0.789 | 3.54 (-10.76-17.85) | 0.718 |
| **First health care provider** | Private outpatient clinic | - |  | - |  |
|  | Public outpatient clinic | 3 (-0.83-6.83) | 0.970 | -1.01 (-10.11-8.08) | 0.931 |
|  | Public Hospital | 0.01 (-1.56-1.56) | 0.883 | -0.19 (-1.86-1.46) | 0.947 |
|  | Private Hospital or nursing home | 0.01 (-1.52-1.52) | 0.887 | 0.71 (-0.50-1.92) | 0.949 |
| **TB diagnosis place** | Private outpatient clinic | - |  |  |  |
|  | Public Hospital | 3 (-8.95-14.95) | 0.791 | 4.38 (-8.16-16.93) | 0.526 |
|  | Private hospital or nursing home | -1.01(-2.01- 0.001) | 0.030 | -0.25 (-1.06-0.54) | 0.494 |

**Table L: Median regression: factors associated with Total delay in Patna.**

| **Characteristics** |  | **Unadjusted** | | **Adjusted** | |
| --- | --- | --- | --- | --- | --- |
|  |  | **Coefficient (95% CI)** | **p-value** | **Coefficient (95% CI)** | **p-value** |
| **Age** |  | 0.188 (-0.340, 0.715) | 0.486 | -0.240 (-1.891, 1.411) | 0.777 |
| **Gender** | Female |  |  |  |  |
|  | Male | -1 (-16.868, 14.868) | 0.902 | 2.403 (-52.229, 57.035) | 0.931 |
| **Marital Status** | Single |  |  |  |  |
|  | Married | 3 (-17.078, 23.078) | 0.77 | 8.910 (-42.781, 60.600) | 0.736 |
|  | Other | 15 (-11.213, 41.213) | 0.263 | 20.768 (-59.656, 101.193) | 0.614 |
| **Number of individuals in the family** |  | 0.273 (-2.271, 2.816) | 0.834 | -2.000 (-9.558, 5.568) | 0.606 |
| **Highest education level** | Primary 6 or lower |  |  |  |  |
|  | Junior secondary school | 8 (-20.260, 36.260) | 0.58 | 14.318 (-37.875, 66.510) | 0.592 |
|  | Senior secondary school | 10 (-36.353, 56.353) | 0.673 | 13.930 (-36.899, 63.636) | 0.743 |
|  | Bachelor's degree | -6 (-32.927, 20.927) | 0.663 | -5.270 (-74.453, 63.913) | 0.882 |
|  | Post-graduate certificate | -13 (-51.649, 25.649) | 0.511 | -34.880 (-141.736, 71.976) | 0.524 |
|  | Other | -9 (-31.402, 13.402) | 0.432 | 13.368 (-69.222, 97.082) | 0.603 |
| **Employment Status** | Working |  |  |  |  |
|  | Self-employed | 0 (-33.783, 33.783) | 1 | 5.742 (-69.073, 80.558) | 0.881 |
|  | Unemployed/underemployed | 1 (-18.047, 20.047) | 0.918 | -6.479 (-58.191, 45.233) | 0.806 |
|  | Retired | -9 (-52.696, 34.696) | 0.687 | 38.526 (-69.250, 146.302) | 0.485 |
| **Financial Standing** | Adequate |  |  |  |  |
|  | Barely adequate | 6 (-12.315, 24.315) | 0.522 | 1.151 (-46.970, 49.273) | 0.963 |
|  | Inadequate | 27 (-4.047, 58.047) | 0.09 | 5.424 (-56.278, 67.126) | 0.864 |
| **Contact with active Pulmonary TB people** | No |  |  |  |  |
|  | Yes | 1 (-14.805, 16.805) | 0.901 | -4.529 (-41.025, 31.966) | 0.808 |
| **Friends or Family COVID-diagnosed** | No |  |  |  |  |
|  | Yes | -11 (-43.109, 21.109) | 0.503 | -11.292 (-109.190, 86.605) | 0.822 |
| **COVID affected willingness to seek healthcare** | No effect |  |  |  |  |
|  | More willing | -15 (-30.658, 0.658) | 0.062 | 14.638 (-34.409, 63.685) | 0.56 |
|  | Less willing | 43 (-41.120, 127.120) | 0.318 | 64.203 (-20.404, 148.810) | 0.14 |
| **First health care provider** | Private outpatient clinic |  |  |  |  |
|  | Public outpatient clinic | 4 (-11.831, 19.831) | 0.621 | -12.289 (-101.531, 76.954) | 0.788 |
|  | Public Hospital | -26 (-49.839, -2.161) | 0.034 | -58.679 (-134.808, 17.451) | 0.134 |
|  | Private Hospital or nursing home | 22 (-8.657, 52.657) | 0.162 | -19.582 (-110.323, 71.158) | 0.673 |
| **TB diagnosis place** | Private outpatient clinic |  |  |  |  |
|  | Public Hospital | -2 (-23.830, 19.830) | 0.858 | 22.254 (-51.515, 96.023) | 0.556 |
|  | Private hospital or nursing home | -7 (-47.509, 33.509) | 0.735 | -1.588 (-90.614, 87.438) | 0.972 |

**Table M: Median regression: factors associated with Total delay in Mumbai.**

|  |  | Unadjusted | | Adjusted | |
| --- | --- | --- | --- | --- | --- |
| Outcome: Total delay |  | **Coefficient (95% CI)** | **p-value** | **Coefficient (95% CI)** | **p-value** |
| Age |  | -0.08 ( -0.59, 0.43) | 0.764 | 0.36 ( -0.55, 1.28) | 0.371 |
| Gender | Female |  |  |  |  |
|  | Male | -6 ( -16.87, 4.87) | 0.281 | -1.62 (-17.89, 14.66) | 0.829 |
| Marital Status | Single |  |  |  |  |
|  | Married | -6 ( -18.79, 6.79) | 0.359 | -12.01 ( -28.25, 4.23) | 0.134 |
|  | Other | -14 ( -85.02, 57.02) | 0.701 | -22.79 ( -87.69, 42.12) | 0.541 |
| Number of individuals in the family |  | 0.20 ( -2.07, 2.47) | 0.863 | -0.30 ( -3.08, 2.48) | 0.826 |
| Highest education level | Primary 6 or lower |  |  |  |  |
|  | Junior secondary school | -0.01 ( -16.17 , 16.17 ) | 0.998 | -2.91 ( -25.06 , 19.24 ) | 0.803 |
|  | Senior secondary school | 1 ( -14.85 , 16.85 ) | 0.902 | 3.34 ( -18.03 , 24.70 ) | 0.772 |
|  | Bachelor's degree | 24 ( -10.88 , 58.88 ) | 0.179 | -1.20 ( -45.00 , 42.61 ) | 0.954 |
|  | Post-graduate certificate | 19 ( -43.39 , 81.39 ) | 0.551 | 22.19 ( -47.34 , 91.72 ) | 0.569 |
|  | Other | 3 ( -11.68 , 17.68 ) | 0.689 | 5.25 ( -17.65 , 28.16 ) | 0.645 |
| Employment Status | Working |  |  |  |  |
|  | Self-employed | 25 ( -33.82 , 83.82 ) | 0.406 | 27.48 ( -35.09 , 90.06 ) | 0.375 |
|  | Unemployed/underemployed | 3 ( -9.43 , 15.43 ) | 0.637 | 2.17 ( -11.12 , 15.45 ) | 0.777 |
|  | Retired | 17 ( -1.59 , 35.59 ) | 0.075 | -35.44 ( -145.06 , 74.18 ) | 0.418 |
| Financial Standing | Adequate |  |  |  |  |
|  | Barely adequate | 1 ( -10.05 , 12.05 ) | 0.859 | 3.08 ( -11.91 , 18.06 ) | 0.715 |
|  | Inadequate | -0.01 ( -51.15 , 51.15 ) | 0.968 | 11.48 ( -29.49 , 52.46 ) | 0.543 |
| Contact with active Pulmonary TB people | No |  |  |  |  |
|  | Yes | 5 ( -14.88, 24.88 ) | 0.623 | 0.84 ( -22.13, 23.80 ) | 0.946 |
| Friends or Family COVID-diagnosed | No |  |  |  |  |
|  | Yes | 3 ( -16.26, 22.26 ) | 0.760 | -6.24 ( -32.99, 20.51 ) | 0.641 |
| COVID affected willingness to seek healthcare | No effect |  |  |  |  |
|  | More willing | -11 ( -22.21, 0.21 ) | 0.056 | -4.07 ( -18.76, 10.61 ) | 0.606 |
|  | Less willing | 9 ( -19.68, 37.68 ) | 0.539 | 38.33 ( -67.65, 144.30 ) | 0.401 |
| First health care provider | Private outpatient clinic |  |  |  |  |
|  | Public outpatient clinic | -11 ( -36.22, 14.22 ) | 0.394 | -0.65 ( -23.21, 21.91 ) | 0.957 |
|  | Public Hospital | -11 ( -23.12, 1.12 ) | 0.077 | -13.96 ( -40.66, 12.74 ) | 0.213 |
|  | Private Hospital or nursing home | -17 ( -34.56, 0.56 ) | 0.059 | -32.45 ( -105.73, 40.82 ) | 0.352 |
| TB diagnosis place | Private outpatient clinic |  |  |  |  |
|  | Public Hospital | 13 ( 5.10, 20.90 ) | 0.001 | 10.25 ( -2.00, 22.51 ) | 0.177 |
|  | Private hospital or nursing home | 120 ( -1.33, 241.33 ) | 0.054 | 34.89 ( -57.48, 127.25 ) | 0.451 |
